# Resting Heart Rate for Global Health Surveillance: Descriptive Epidemiology of 566,986 Participants from the WHO STEP Surveys and the South Asia Biobank

**DOI:** 10.64898/2026.11.11.26367828

**Authors:** Abdulwahab Alshallal, Tomas Gonzales, Antony Siahaan, Wubin Xie, Louise Foley, Parizad Avari, Guha Pradeepa, Sara Mahmood, Awais Butt, Laksara De Silva, Madawa Chandrathilake, Akansha Tyagi, Menka Loomba, Mehedi Hasan, Ali Ahsan, Tanmoy Sarker, Marie Loh, Sajjad Ahmed, Anuradhani Kasturiratne, Malay Mridha, R.M. Anjana, Prasad Katulanda, Vinitaa Jha, Khadija I. Khawaja, John C. Chambers, Soren Brage, WHO Country Author Group, South Asia Biobank Study Group

## Abstract

**Objective:** To describe global patterns of resting heart rate (RHR) in population subgroups and over time.

**Methods:** Observational data from WHO STEP surveys (n=363,462 participants from 76 countries, 2002-2020) and the South Asia Biobank (SAB, n=203,524 participants from four South Asian countries, 2018-2023). RHR was measured in triplicate while seated using a blood pressure cuff. Height and weight were measured using anthropometric techniques. Physical activity was self-reported and accelerometry-measured (SAB subsample, n=22,183). We describe RHR distributions by geographical region, temporality, sex, age, BMI, and physical activity. Utility of RHR to explain accelerometer-based physical activity was examined by regression.

**Results:** Country-level median RHR ranged 65–80 bpm, higher in women than men. Age-related RHR patterns diverged markedly by sex; men showing a curvilinear association with higher RHR in midlife, and women showing a consistent decline with age across most regions. Compared to normal-weight, obese participants had higher RHR [WHO: (men: +4.4 bpm, women: +0.9 bpm, SAB: (men: +3.1 bpm, women: +1.0 bpm)]. Participants meeting physical activity recommendations had lower RHR [WHO: (men –1.6 bpm, women: −0.9 bpm), SAB: (men: −2.1 bpm, women: −0.5 bpm)] compared to insufficiently active. In SAB (field-work spanning whole-calendar year), seasonal variations were present (0.6–0.9 bpm higher RHR in June-August compared to the Sept-Nov period), most pronounced further from the equator. In 21 WHO countries with repeat survey data, RHR was stable over time in women but declined in men (−0.4 bpm/year). Self-reported activity explained 2-14% (women: 2-3%, men: 10-14%) of the variance in device-measured physical activity, and RHR added 1-3% incrementally.

**Conclusions:** RHR varies globally, exhibits different age-related patterns by sex, and is higher in population subgroups with greater non-communicable disease burden. RHR is easy to measure and may have utility for global health surveillance.

## INTRODUCTION

Cardiorespiratory fitness (CRF) is a strong predictor of morbidity and mortality (1). Low CRF is linked to obesity which affects over 1 billion people globally (2). Despite both fitness and obesity being modifiable risk factors, there is currently no global health surveillance system in place to monitor fitness, with levels being only sporadically described in population studies (3). A small proportion of studies have described CRF in European populations, Latin America, Africa, and South Asia (4–7). In Africa, different ethnic populations in Kenya, South Africa, and Tanzania exhibit large differences in CRF (7–9). Of the limited research available, it has been shown that CRF tends to be higher in men than women and declines with age (9–11). South Asian men living in the UK have lower CRF compared to age- and BMI-matched white men (12).

The gold-standard measurement of CRF is maximal oxygen consumption (VO_2_max) during an incremental exercise test to exhaustion. The feasibility of this assessment is, however, challenging in most situations, making it difficult to incorporate into surveillance systems. Therefore, alternative methods are typically deployed, including heart rate response to submaximal exercise tests (13) but most cohort studies do not include dynamic exercise testing. The most feasible indicator of CRF is resting heart rate (RHR) which is widely measured across national surveys, cohort studies and clinical settings; RHR has been shown to be inversely correlated with CRF (14). Additionally, individuals with low CRF who have metabolic disease or are at an elevated risk of metabolic disease have significantly higher RHR compared to healthy individuals (15,16). RHR is also positively associated with cancer and CVD-related mortality (17,18).

To gain insights into the global variation of RHR and its potential for health surveillance, we describe the global epidemiology of RHR, with an enhanced emphasis on South Asia that has relatively high levels of NCDs. Specifically, we integrate data from the WHO STEP surveys which collected RHR data from 76 countries globally, with the South Asia Biobank (SAB) which is a multicenter study focused on chronic diseases in four South Asian countries with additional objective measures of lung function and device-based physical activity (19,20). This dual-dataset approach provides a broad global overview while offering fine-grained insights into South Asia, allowing for comparisons of RHR levels between populations and assessing its potential utility for routine surveillance of cardiorespiratory health.

## METHODS

### Study Populations

#### WHO STEP Surveys

The WHO STEPS surveys, first administered in 2002, follow the WHO STEP-wise approach to noncommunicable disease risk factor surveillance (19). This analysis includes data on representative samples of 363,462 individuals aged 18 and above from 76 countries (100 unique surveys) across 7 world regions. In total, 21 countries conducted repeat surveys (Micronesia: four surveys; Benin: three, the remaining 19: two each). Step 1 of the survey is questionnaire-based and includes demographic variables and behavioral information on diet, physical activity, smoking, alcohol consumption, and disease status. Step 2 focuses on physical measurements, including anthropometry, resting blood pressure and heart rate.

#### South Asia Biobank

SAB is a large-scale multi-center cohort study investigating determinants of chronic diseases in South Asian populations (20). Between 2018 and 2023, the SAB study recruited 205,356 adults aged 18 years and above, from 564 surveillance sites across four countries or five test regions (Bangladesh, North India, South India, Pakistan, and Sri Lanka). At the baseline visit, participants attended local study assessment centers in the fasting state, for a single visit, during which they completed a structured assessment that was standardized across all settings. Data include self-report (demographics, diet, physical activity, smoking, alcohol, disease status), physical measurements (anthropometry, RHR, blood pressure, lung function), and device-based physical activity measures (accelerometry) in a subsample.

### Measurements

#### Resting Heart Rate

RHR was measured as the pulse rate during seated rest when blood pressure was assessed using automated sphygmomanometers. Measurements were performed in triplicate in Step 2 in WHO STEPS and during the baseline clinical visit in SAB. Values below 30 bpm and above 150 bpm were discarded as invalid. The minimum pulse rate across valid measures for each individual was used as RHR in the present analyses to reflect the most rested state. We also conducted a sensitivity analysis using the median value.

#### Stratifying variables

RHR was described in different population subgroups: Sex (male, female), age group (18-24, 25-44, 45-64, 65+), BMI groups (underweight [<18.5, normal [18.5-24.9], overweight [25-29.9], obese[30+]), self-reported moderate-to-vigorous physical activity (MVPA) as assessed by the global physical activity questionnaire (meeting/not meeting 150 min/week MVPA guideline), dietary intake (consume five or more servings of fruits and vegetables daily/ does not consume five or more servings of fruits and vegetables daily/ missing), smoking status (smoker/non-smoker), alcohol status (drinker/non-drinker), marital status (single/married or cohabiting/divorced or separated/widowed), occupation (government/non-government employee/self-employed/ homemaker/unemployed/ Student/Retired/Non-paid/Missing), and education (no formal schooling, less than primary school completed, primary school completed, secondary school completed, college, postgraduate, missing). Levels for some covariates (e.g., occupation and education) differed between datasets due to survey-specific categorisations but were harmonised for analysis where possible.

In WHO STEPS, countries were also aggregated into regions (Sub-Saharan Africa, Central and East Asia, East Europe and West Asia, Latin and South America, Middle East North Africa, Small Island Nations, South and Southeast Asia) to describe broader place variation. Time (era) was parameterised as survey year centered on 2010. Income was parameterised using country-specific classification (low, lower-middle, upper-middle, high).

In SAB, test region (Pakistan, North India, South India, Sri Lanka, and Bangladesh) was used to explore place differences. Time was expressed as calendar year centered on 2021 (mid-survey year) to capture any secular trends over the short time span of this fieldwork, whereas seasonal variation within year was captured as month of year. Personal income was parameterised in country-specific quartiles. Prevalent metabolic disease status (yes/no) was concatenated from clinical interview questions of hypertension, hypercholesterolemia, and type 2 diabetes. Fat mass was derived from bioelectric impedance and expressed relative to height squared as fat mass index (FMI) to provide an alternative adiposity measure to BMI. Physical activity was also measured objectively with a wrist-worn accelerometer in a subsample (n=22,183); MVPA was defined as wrist acceleration above 125 mg, aggregated in 5-sec analysis epochs and expressed as average weekly MVPA duration (min/week). Lung function, another proxy indicator of cardiorespiratory fitness, was measured by spirometry. Forced vital capacity (FVC) and forced expiratory volume (FEV1) were measured in triplicate, and the highest value across reproducible trials (within 0.15L of the second-best value) was retained per participant. FEV/FVC ratio was derived from these and categorised as high (<0.5), medium (0.5 to 0.7), and low (>0.7) obstruction to distinguish levels of airflow obstruction (21).

### Statistical Analysis

SAB and WHO data were analysed separately.

#### Regional differences

Sex-stratified RHR percentiles (5^th^, 25^th^, 50^th^, 75^th^, 95^th^) were calculated and visualized using boxplots, violin plots, and line plots, stratified by population subgroup. The association with age was modelled using cubic splines with three degrees of freedom, fitted as a three-way interaction with sex and world region. In SAB, the same specification was applied but with test region to examine patterns between SAB test regions. Place was examined by country and world region. The BMI-RHR association was examined ecologically to assess if RHR may rank countries by this NCD risk factor.

To align WHO and SAB results, common analytical methods were applied where possible, with extensions for more detailed information in SAB. To examine influence of survey approach, RHR distributions were compared in countries (Bangladesh and Sri Lanka) which had data from both WHO and SAB using medians (with percentiles), violin plots, boxplots, and Wilcoxon rank-sum tests, stratified by sex, age group, and survey.

#### Population subgroups

BMI was examined both categorically (underweight, normal, overweight, obese) within the multilevel models described below, and continuously. For the continuous analysis, cubic splines were fitted separately within each sex, adjusting for country (WHO) or test region (SAB). For SAB, additional associations of RHR with FMI, lung function categories and device-based physical activity tertiles were explored. Potential regional heterogeneity in associations was explored by region-stratified analyses.

#### Time-related variation in RHR

In WHO STEPS, population median RHR trends over time (survey year centered on 2010) were visualized via line plots in the 19 countries with repeat surveys; a subset regression model (countries with repeats) specifically assessed secular trends. In SAB, seasonal variation in RHR was examined by test region using both cosinor models and time-of-year categories (Sept–Nov, Dec–Feb, Mar–May, June–Aug), with adjustment for calendar year (centered on 2021) to capture any short-term secular patterns.

#### Multivariable regression analyses

We examined RHR patterns in relation to all above-considered explanatory variables in sex-stratified regression models. We used generalized linear multilevel models with a Gamma distribution and log link mutually adjusted for all explanatory variables, levels specified as individual and country to account for clustering by country. In WHO, analyses were weighted by population size. As SAB is a community-based study not necessarily designed for national representativeness, unweighted analyses were used.

#### Utility of resting heart rate to routine physical activity surveillance

In SAB, crude linear regression models evaluated RHR as a potential additional predictor of device-based physical activity, comparing models with GPAQ alone and RHR + GPAQ. For completeness, we repeated this evaluation swapping GPAQ as the dependent variable and treating device-based MVPA as predictor. Model performance was evaluated via R^2^ values. Analyses were stratified by sex and age group (18–44 and 45+ years) and additionally by country, to explore RHR’s complementary value to self-reported or device-based physical activity measures in population subgroups.

## RESULTS

A total of 363,462 and 203,524 participants are included in the present analysis for WHO and SAB, respectively (flowchart, supplementary figure S1). In WHO STEPS, included participants were slightly older, less obese, more likely to be married, and less likely to smoke compared to participants excluded due to missing data. In SAB, included participants were more obese, more likely to be married or cohabiting, and exhibited higher smoking rates (men only) than excluded participants (Supplementary material S1).

Women had higher RHR (WHO: 76 bpm, SAB: 79) than men (WHO: 72 bpm, SAB: 70). Expanded results are in the appendix (Supplementary Material S2). Sex-stratified multivariable regression analyses were conducted for each dataset (Tables 1 and 2). Using the median RHR instead of the minimum RHR of 3 readings did not materially change associations for any of the analyses (data not shown).

**Table 1:** Sex-stratified univariate and multivariable associations for sociodemographic predictors of resting heart rate in WHO STEPS.

|  | Men |  |  |  | Women |  |  |  |
|---|---|---|---|---|---|---|---|---|
|  | Univariate |  | Mutually Adjusted |  | Univariate |  | Mutually Adjusted |  |
| Variable | Estimate | 95% CI | Estimate | 95% CI | Estimate | 95% CI | Estimate | 95% CI |
| **Age Group** |  |  |  |  |  |  |  |  |
| 18-24 | 0.978 | (0.971, 0.986) | 0.989 | (0.980, 0.998) | 1.018 | (1.012, 1.024) | 1.020 | (1.014, 1.026) |
| 25-44 | *Ref* | – | *Ref* | – | *Ref* | – | *Ref* | – |
| 45-64 | 1.019 | (1.014, 1.024) | 1.018 | (1.013, 1.023) | 0.985 | (0.982, 0.989) | 0.983 | (0.979, 0.987) |
| 65+ | 1.021 | (1.012, 1.031) | 1.015 | (1.005, 1.025) | 0.991 | (0.982, 1.000) | 0.987 | (0.978, 0.997) |
| **BMI Group** |  |  |  |  |  |  |  |  |
| Underweight | 1.013 | (1.007, 1.019) | 1.008 | (1.002, 1.014) | 1.008 | (1.003, 1.014) | 1.009 | (1.003, 1.014) |
| Normal | *Ref* | – | *Ref* | – | *Ref* | – | *Ref* | – |
| Overweight | 1.030 | (1.024, 1.036) | 1.030 | (1.023, 1.036) | 1.005 | (1.001, 1.009) | 1.007 | (1.003, 1.012) |
| Obese | 1.061 | (1.051, 1.071) | 1.060 | (1.049, 1.070) | 1.013 | (1.007, 1.018) | 1.017 | (1.012, 1.023) |
| **Physical Activity Level** |  |  |  |  |  |  |  |  |
| Less than 150 min/week | *Ref* | – | *Ref* | – | *Ref* | – | *Ref* | – |
| 150+ min/week | 0.977 | (0.972, 0.983) | 0.986 | (0.981, 0.992) | 0.995 | (0.991, 0.999) | 0.994 | (0.990, 0.998) |
| **Daily Consumption of Fruits and Vegetables** |  |  |  |  |  |  |  |  |
| Less than 5 servings | *Ref* | – | *Ref* | – | *Ref* | – | *Ref* | – |
| At Least 5 Servings | 0.998 | (0.990, 1.006) | 0.999 | (0.991, 1.007) | 0.993 | (0.987, 0.999) | 0.994 | (0.987, 1.000) |
| Missing Diet | 0.996 | (0.991, 1.001) | 1.000 | (0.995, 1.005) | 0.996 | (0.993, 1.000) | 0.997 | (0.993, 1.000) |
| **Smoking Status** |  |  |  |  |  |  |  |  |
| Non-Smoker | *Ref* | – | *Ref* | – | *Ref* | – | *Ref* | – |
| Smoker | 1.007 | (1.002, 1.011) | 1.011 | (1.006, 1.016) | 0.998 | (0.988, 1.007) | 1.003 | (0.993, 1.012) |
| Missing Smoking Status | 0.987 | (0.953, 1.022) | 0.957 | (0.924, 0.991) | 1.022 | (1.004, 1.040) | 1.029 | (1.010, 1.048) |
| **Alcohol Drinking Status** |  |  |  |  |  |  |  |  |
| Non-Drinker | *Ref* | – | *Ref* | – | *Ref* | – | *Ref* | – |
| Drinker | 1.003 | (0.996, 1.010) | 1.011 | (1.003, 1.018) | 0.985 | (0.979, 0.992) | 0.992 | (0.985, 0.999) |
| Missing Drinking Status | 1.013 | (1.007, 1.020) | 1.024 | (1.017, 1.031) | 0.987 | (0.983, 0.992) | 0.994 | (0.987, 1.000) |
| **Marital Status** |  |  |  |  |  |  |  |  |
| Married or Cohabiting | *Ref* | – | *Ref* | – | *Ref* | – | *Ref* | – |
| Never Married | 0.987 | (0.980, 0.994) | 1.006 | (0.997, 1.014) | 1.011 | (1.004, 1.018) | 1.006 | (0.999, 1.014) |
| Separated/Divorced | 1.026 | (1.009, 1.043) | 1.027 | (1.010, 1.044) | 0.999 | (0.991, 1.007) | 1.003 | (0.995, 1.011) |
| Widowed | 1.027 | (1.006, 1.048) | 1.018 | (0.998, 1.040) | 0.993 | (0.986, 0.999) | 1.001 | (0.994, 1.007) |
| Missing Marital Status | 1.028 | (1.023, 1.034) | 1.000 | (0.991, 1.009) | 1.025 | (1.020, 1.029) | 1.031 | (1.024, 1.039) |
| **Highest Education Level** |  |  |  |  |  |  |  |  |
| No formal schooling | *Ref* | – | *Ref* | – | *Ref* | – | *Ref* | – |
| College/University completed | 1.006 | (0.996, 1.015) | 1.003 | (0.992, 1.013) | 0.992 | (0.984, 1.001) | 0.987 | (0.978, 0.997) |
| Less than primary school | 0.999 | (0.992, 1.006) | 1.002 | (0.996, 1.009) | 1.001 | (0.996, 1.006) | 0.996 | (0.991, 1.001) |
| Post graduate degree | 1.015 | (1.001, 1.028) | 1.008 | (0.994, 1.023) | 0.988 | (0.976, 1.001) | 0.988 | (0.974, 1.001) |
| Primary school completed | 0.997 | (0.991, 1.004) | 1.005 | (0.999, 1.012) | 1.008 | (1.004, 1.013) | 1.001 | (0.995, 1.006) |
| Secondary school completed | 1.006 | (0.999, 1.013) | 1.007 | (0.999, 1.014) | 1.005 | (0.999, 1.010) | 0.997 | (0.991, 1.003) |
| Missing Education | 1.010 | (1.003, 1.017) | 1.008 | (1.000, 1.016) | 1.001 | (0.996, 1.007) | 0.993 | (0.987, 0.999) |
| **Occupation** |  |  |  |  |  |  |  |  |
| Self-employed | *Ref* | – | *Ref* | – | *Ref* | – | *Ref* | – |
| Government employee | 1.009 | (1.001, 1.018) | 1.003 | (0.994, 1.011) | 0.996 | (0.987, 1.005) | 0.999 | (0.989, 1.009) |
| Homemaker | 0.990 | (0.979, 1.000) | 0.989 | (0.979, 0.999) | 1.009 | (1.003, 1.015) | 1.009 | (1.003, 1.015) |
| Non-government employee | 1.005 | (0.998, 1.012) | 1.005 | (0.997, 1.012) | 0.999 | (0.991, 1.008) | 0.995 | (0.986, 1.003) |
| Non-paid | 1.019 | (1.006, 1.031) | 1.016 | (1.003, 1.028) | 1.007 | (0.994, 1.021) | 1.007 | (0.993, 1.020) |
| Retired | 0.997 | (0.985, 1.010) | 0.995 | (0.983, 1.008) | 0.995 | (0.982, 1.008) | 0.993 | (0.980, 1.006) |
| Student | 0.991 | (0.984, 0.997) | 0.991 | (0.985, 0.998) | 0.997 | (0.989, 1.006) | 0.995 | (0.986, 1.003) |
| Unemployed | 0.989 | (0.982, 0.996) | 0.993 | (0.985, 1.000) | 1.014 | (1.007, 1.020) | 1.014 | (1.006, 1.021) |
| Missing Occupation | 1.015 | (1.007, 1.023) | 1.008 | (1.000, 1.017) | 1.029 | (1.024, 1.035) | 1.025 | (1.018, 1.031) |
| **Country Income Class** |  |  |  |  |  |  |  |  |
| Low | 0.989 | (0.984, 0.994) | 1.034 | (1.027, 1.042) | 0.999 | (0.995, 1.003) | 1.015 | (1.010, 1.021) |
| Lower-middle | *Ref* | – | *Ref* | – | *Ref* | – | *Ref* | – |
| Upper-middle | 1.009 | (0.997, 1.021) | 0.998 | (0.983, 1.013) | 0.967 | (0.958, 0.975) | 1.001 | (0.990, 1.012) |
| High | 1.027 | (0.989, 1.066) | 1.000 | (0.963, 1.039) | 0.992 | (0.965, 1.020) | 1.010 | (0.982, 1.039) |
| **Region** |  |  |  |  |  |  |  |  |
| Sub-Saharan Africa | *Ref* | – | *Ref* | – | *Ref* | – | *Ref* | – |
| MENA | 1.026 | (1.019, 1.034) | 1.036 | (1.027, 1.045) | 1.007 | (1.002, 1.013) | 1.018 | (1.011, 1.025) |
| East Europe and West Asia | 1.034 | (1.020, 1.048) | 1.063 | (1.045, 1.081) | 0.952 | (0.944, 0.961) | 0.983 | (0.970, 0.996) |
| Central and East Asia | 1.058 | (1.037, 1.080) | 1.080 | (1.057, 1.104) | 0.975 | (0.960, 0.990) | 1.005 | (0.988, 1.022) |
| South and South-East Asia | 1.030 | (1.025, 1.035) | 1.048 | (1.040, 1.055) | 1.005 | (1.002, 1.009) | 1.017 | (1.010, 1.023) |
| Small Island Nations | 0.963 | (0.893, 1.037) | 0.967 | (0.898, 1.042) | 0.942 | (0.888, 0.999) | 0.945 | (0.891, 1.003) |
| Latin and South America | 1.008 | (0.990, 1.026) | 1.049 | (1.027, 1.071) | 0.970 | (0.957, 0.984) | 0.987 | (0.971, 1.004) |
| Year (relative to 2010) | 0.994 | (0.994, 0.995) | 0.994 | (0.993, 0.995) | 0.999 | (0.999, 1.000) | 1.000 | (1.000, 1.001) |

Coefficients represent multiplicative difference in RHR relative to the reference category.
Mutually adjusted models are adjusted for all explanatory variables in the table.
Full results are in appendix.

**Table 2:** Sex-stratified univariate and multivariable associations for sociodemographic predictors of resting heart rate in SAB.

|  | Men |  |  |  | Women |  |  |  |
|---|---|---|---|---|---|---|---|---|
|  | Univariate |  | Mutually Adjusted |  | Univariate |  | Mutually Adjusted |  |
| Variable | Estimate | 95% CI | Estimate | 95% CI | Estimate | 95% CI | Estimate | 95% CI |
| **Age Group** |  |  |  |  |  |  |  |  |
| 18-24 | 1.030 | (1.024, 1.036) | 1.002 | (0.996, 1.008) | 1.053 | (1.049, 1.057) | 1.043 | (1.039, 1.048) |
| 25-44 | *Ref* | – | *Ref* | – | *Ref* | – | *Ref* | – |
| 45-64 | 1.011 | (1.009, 1.014) | 0.998 | (0.996, 1.001) | 0.973 | (0.971, 0.975) | 0.965 | (0.963, 0.967) |
| 65+ | 0.989 | (0.985, 0.993) | 0.979 | (0.976, 0.983) | 0.961 | (0.958, 0.964) | 0.955 | (0.952, 0.958) |
| **BMI Group** |  |  |  |  |  |  |  |  |
| Underweight | 1.007 | (1.003, 1.011) | 1.023 | (1.019, 1.027) | 1.011 | (1.007, 1.015) | 1.012 | (1.008, 1.015) |
| Normal | *Ref* | – | *Ref* | – | *Ref* | – | *Ref* | – |
| Overweight | 1.065 | (1.062, 1.068) | 1.018 | (1.015, 1.020) | 1.018 | (1.016, 1.020) | 1.002 | (1.000, 1.004) |
| Obese | 1.127 | (1.122, 1.132) | 1.033 | (1.029, 1.038) | 1.058 | (1.055, 1.060) | 1.011 | (1.009, 1.014) |
| **Disease History** |  |  |  |  |  |  |  |  |
| No | *Ref* | – | *Ref* | – | *Ref* | – | *Ref* | – |
| Yes | 1.051 | (1.049, 1.054) | 1.038 | (1.036, 1.041) | 1.016 | (1.015, 1.018) | 1.026 | (1.024, 1.028) |
| Missing | 1.034 | (1.022, 1.047) | 0.965 | (0.942, 0.989) | 1.010 | (1.001, 1.019) | 1.004 | (0.984, 1.024) |
| **Physical Activity Level** |  |  |  |  |  |  |  |  |
| Less than 150 min/week | *Ref* | – | *Ref* | – | *Ref* | – | *Ref* | – |
| At least 150 min/week | 0.902 | (0.900, 0.905) | 0.979 | (0.976, 0.982) | 0.943 | (0.941, 0.945) | 0.990 | (0.988, 0.992) |
| **Daily Consumption of Fruits and Vegetables** |  |  |  |  |  |  |  |  |
| Less than 5 Servings | *Ref* | – | *Ref* | – | *Ref* | – | *Ref* | – |
| At least 5 Servings | 0.979 | (0.976, 0.983) | 0.998 | (0.994, 1.001) | 0.993 | (0.990, 0.996) | 1.002 | (1.000, 1.005) |
| Missing | 0.962 | (0.959, 0.965) | 0.994 | (0.991, 0.997) | 0.995 | (0.993, 0.997) | 0.998 | (0.996, 1.000) |
| **Smoking Status** |  |  |  |  |  |  |  |  |
| Non-Smoker | *Ref* | – | *Ref* | – | *Ref* | – | *Ref* | – |
| Smoker | 0.939 | (0.936, 0.941) | 0.979 | (0.976, 0.981) | 0.976 | (0.963, 0.989) | 0.970 | (0.958, 0.982) |
| Missing | 1.037 | (1.023, 1.051) | – | – | 1.017 | (1.007, 1.026) | – | – |
| **Alcohol Drinking Status** |  |  |  |  |  |  |  |  |
| Never drank | *Ref* | – | *Ref* | – | *Ref* | – | *Ref* | – |
| Frequent | 1.064 | (1.056, 1.071) | 1.039 | (1.032, 1.046) | 0.953 | (0.915, 0.992) | 0.985 | (0.948, 1.023) |
| Infrequent | 0.975 | (0.970, 0.980) | 0.996 | (0.991, 1.001) | 0.927 | (0.914, 0.939) | 0.953 | (0.941, 0.966) |
| Moderate | 1.003 | (0.997, 1.010) | 1.002 | (0.996, 1.008) | 0.941 | (0.912, 0.971) | 0.965 | (0.938, 0.994) |
| Past | 0.970 | (0.966, 0.974) | 1.005 | (1.001, 1.008) | 1.021 | (1.012, 1.029) | 0.993 | (0.985, 1.000) |
| Missing | 1.050 | (1.036, 1.064) | – | – | 1.017 | (1.007, 1.026) | – | – |
| **Marital Status** |  |  |  |  |  |  |  |  |
| Married or Cohabiting | *Ref* | – | *Ref* | – | *Ref* | – | *Ref* | – |
| Divorced/Separated | 1.061 | (1.043, 1.080) | 1.014 | (0.999, 1.029) | 0.982 | (0.974, 0.990) | 0.992 | (0.984, 0.999) |
| Never married | 1.035 | (1.030, 1.040) | 1.014 | (1.009, 1.020) | 1.056 | (1.052, 1.061) | 1.011 | (1.006, 1.016) |
| Widowed | 1.034 | (1.023, 1.045) | 1.012 | (1.003, 1.022) | 0.979 | (0.976, 0.982) | 0.995 | (0.992, 0.998) |
| Missing | 1.056 | (1.042, 1.071) | – | – | 1.017 | (1.008, 1.027) | – | – |
| **Highest Education Level** |  |  |  |  |  |  |  |  |
| No Formal Schooling | *Ref* | – | *Ref* | – | *Ref* | – | *Ref* | – |
| High school completed | 1.036 | (1.031, 1.040) | 1.011 | (1.007, 1.016) | 0.998 | (0.995, 1.002) | 1.003 | (1.000, 1.007) |
| Less than primary school | 0.959 | (0.955, 0.963) | 0.997 | (0.993, 1.001) | 0.954 | (0.951, 0.957) | 1.000 | (0.997, 1.003) |
| Primary school completed | 0.977 | (0.973, 0.980) | 1.004 | (1.001, 1.008) | 0.972 | (0.970, 0.975) | 1.009 | (1.006, 1.011) |
| Secondary school completed | 1.009 | (1.005, 1.013) | 1.007 | (1.003, 1.011) | 0.979 | (0.976, 0.982) | 1.007 | (1.004, 1.010) |
| University | 1.036 | (1.027, 1.046) | 1.014 | (1.006, 1.023) | 0.998 | (0.988, 1.007) | 1.000 | (0.991, 1.009) |
| Missing | 1.046 | (1.041, 1.052) | 1.005 | (1.000, 1.011) | 1.003 | (0.999, 1.008) | 0.993 | (0.988, 0.998) |
| **Occupation** |  |  |  |  |  |  |  |  |
| Self-employed | *Ref* | – | *Ref* | – | *Ref* | – | *Ref* | – |
| Agricultural worker | 0.929 | (0.925, 0.933) | 0.976 | (0.972, 0.979) | 0.968 | (0.962, 0.975) | 0.989 | (0.982, 0.995) |
| Cleaner/labourer | 0.989 | (0.984, 0.993) | 0.984 | (0.980, 0.988) | 1.020 | (1.014, 1.026) | 0.986 | (0.981, 0.992) |
| Non-government employee | 1.059 | (1.055, 1.063) | 1.003 | (0.999, 1.006) | 1.005 | (1.000, 1.010) | 0.990 | (0.985, 0.995) |
| Homemaker | 1.129 | (1.120, 1.138) | 1.019 | (1.011, 1.026) | 1.023 | (1.019, 1.027) | 1.009 | (1.005, 1.013) |
| Unemployed | 1.068 | (1.063, 1.073) | 1.023 | (1.019, 1.028) | 1.039 | (1.035, 1.043) | 1.021 | (1.018, 1.025) |
| Other | 1.037 | (1.033, 1.042) | 1.003 | (0.998, 1.007) | 1.023 | (1.017, 1.028) | 1.001 | (0.996, 1.006) |
| **Income Quartile** |  |  |  |  |  |  |  |  |
| Q1 | *Ref* | – | *Ref* | – | *Ref* | – | *Ref* | – |
| Q2 | 1.019 | (1.015, 1.023) | 0.998 | (0.995, 1.002) | 1.014 | (1.012, 1.017) | 1.000 | (0.998, 1.002) |
| Q3 | 1.042 | (1.038, 1.046) | 0.999 | (0.995, 1.002) | 1.002 | (1.000, 1.005) | 0.999 | (0.996, 1.001) |
| Q4 | 1.023 | (1.019, 1.026) | 1.001 | (0.997, 1.005) | 0.985 | (0.983, 0.988) | 1.002 | (0.999, 1.005) |
| Missing | 1.075 | (1.061, 1.090) | 1.063 | (1.034, 1.093) | 1.018 | (1.008, 1.027) | 1.020 | (0.997, 1.044) |
| **Time of Year** |  |  |  |  |  |  |  |  |
| Sept-Nov | *Ref* | – | *Ref* | – | *Ref* | – | *Ref* | – |
| Dec-Feb | 0.973 | (0.970, 0.976) | 0.982 | (0.979, 0.985) | 0.972 | (0.970, 0.974) | 0.982 | (0.980, 0.984) |
| Mar-May | 0.976 | (0.973, 0.980) | 0.998 | (0.995, 1.002) | 0.987 | (0.985, 0.990) | 0.997 | (0.995, 0.999) |
| June-Aug | 1.013 | (1.009, 1.016) | 1.013 | (1.009, 1.016) | 1.005 | (1.003, 1.008) | 1.008 | (1.005, 1.010) |
| Year (relative to 2021) | 0.992 | (0.991, 0.993) | 1.000 | (0.999, 1.001) | 1.003 | (1.003, 1.004) | 1.001 | (1.000, 1.002) |
| **Test Region** |  |  |  |  |  |  |  |  |
| Bangladesh | *Ref* | – | *Ref* | – | *Ref* | – | *Ref* | – |
| India north | 1.178 | (1.173, 1.184) | 1.131 | (1.126, 1.137) | 1.079 | (1.075, 1.082) | 1.077 | (1.073, 1.081) |
| India south | 1.223 | (1.218, 1.228) | 1.179 | (1.173, 1.184) | 1.053 | (1.049, 1.057) | 1.063 | (1.059, 1.067) |
| Pakistan | 1.222 | (1.218, 1.226) | 1.175 | (1.170, 1.179) | 1.140 | (1.138, 1.143) | 1.127 | (1.123, 1.130) |
| Sri Lanka | 1.062 | (1.059, 1.065) | 1.030 | (1.026, 1.034) | 1.019 | (1.017, 1.021) | 1.016 | (1.013, 1.019) |

Estimates represent multiplicative difference in RHR relative to the reference category.
In mutually adjusted models, the “Missing” category was excluded due to perfect collinearity arising from identical missingness patterns across alcohol drinking status, smoking status, and marital status.
Mutually adjusted models are adjusted for all explanatory variables in the table.
Full results are in appendix.

### Age-related patterns and geographic variation

Across geographical regions, RHR exhibited a curvilinear relationship with age in men, rising through early and middle adulthood before declining in older age groups, whereas women showed a consistent decline in RHR with older age (Figure 1). The age at which RHR peaked in men varied by region, but the curvilinear shape was broadly consistent. This sex divergence was evident at the region level but not always at the country level (Figure S2), where between-country variation obscured the pattern. In SAB, the divergence was different in the pooled panel where men showed a rise but then a decline in older ages whereas women showed progressively lower RHR across older age groups. Age remained independently associated with RHR after mutual adjustment for all covariates in both datasets (Tables 1 and 2).

**Figure 1.**
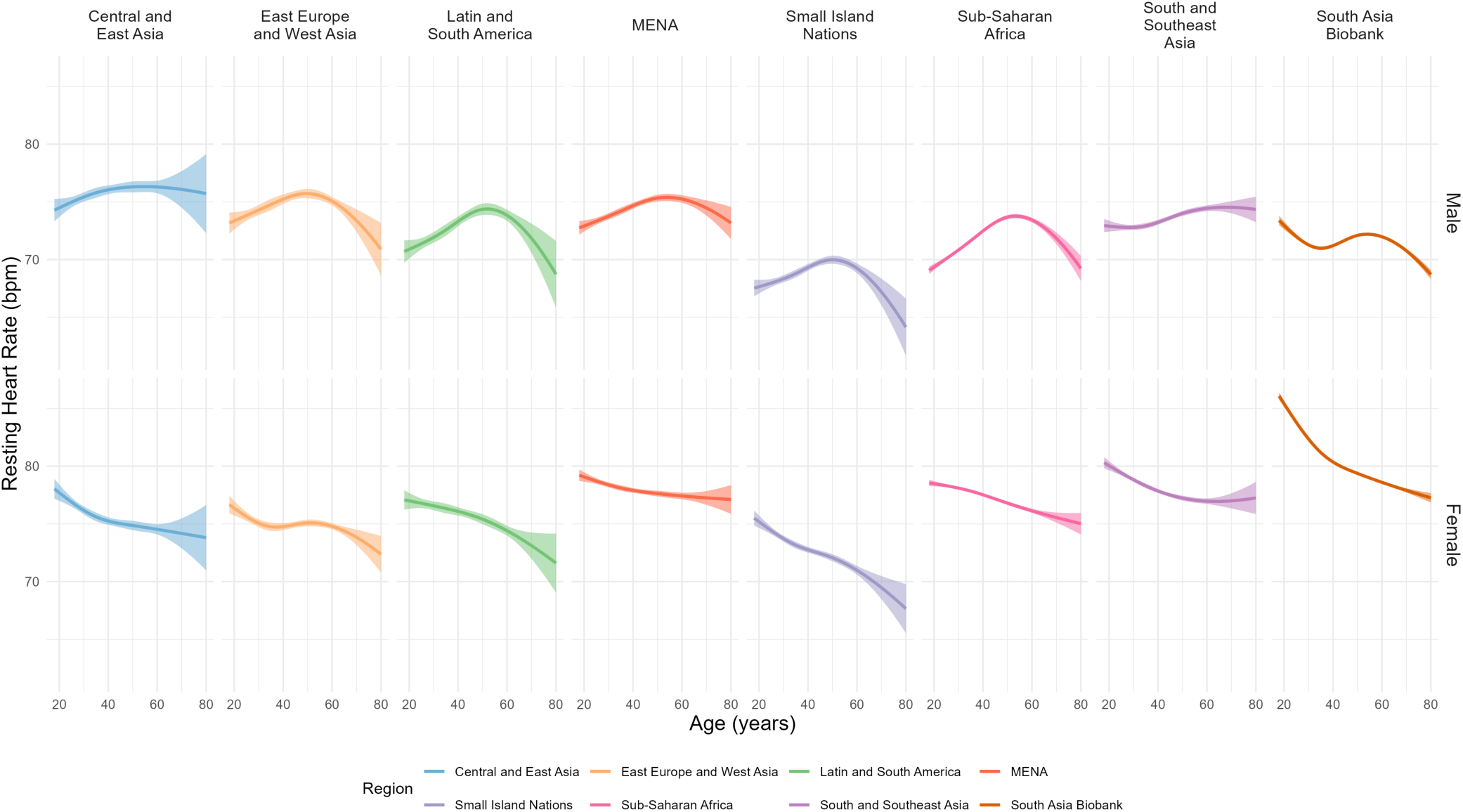
Sex-stratified cubic spline plots for the association between age and resting heart rate by region.

Associations with age varied by SAB test region (Supplementary Figure S3). Men in India showed curvilinear patterns resembling those in WHO regions, with a midlife peak followed by decline. Men in Bangladesh and Sri Lanka showed broadly monotonic declines, and men in Pakistan showed a plateau through midlife before declining. In women, patterns were largely monotonic across all SAB regions, consistent with the WHO regional findings.

The lowest median RHR values were observed in the Small Island Nations region (men: 68 bpm; women: 72 bpm) and the highest in MENA and Sub-Saharan Africa (men: 80 bpm; women: 86 bpm), with some evidence of multimodality; however country-level distributions were not multimodal (Figure S4a). Within SAB, Pakistan had the highest median RHR (men: 80 bpm; women: 86 bpm) and Bangladesh the lowest for both sexes (men: 64 bpm, women: 75 bpm), with intermediate values across the remaining test regions (Figure S4b). In the two countries with data from both surveys (Bangladesh and Sri Lanka), men generally had lower RHR in SAB than in WHO STEPS. Significant differences were found across all age and sex strata (Figure S5). RHR distributions and density plots stratified by age group, sex, and region are shown in Supplementary Figure S6.

### Weight status

The relationship between BMI and RHR was non-linear in most populations. In WHO, cubic spline models fitted separately by sex showed a RHR nadir at a BMI of 20.7 kg/m^2^ in men and 22.2 kg/m^2^ in women; 26.6% and 31.9% of the sample are below these BMI thresholds, respectively. In SAB, RHR nadirs at BMI of 21.0 kg/m^2^ in men and 23.5 kg/m^2^ in women were observed, with 29.3% and 36.9% of the sample, respectively, falling below these (Figure 2). Region-stratified splines showed largely similar results across WHO regions and SAB test regions (Supplementary Figure S7). The category-based multilevel models were broadly consistent with this pattern showing higher RHR in underweight, overweight, and obese participants compared to normal-weight in both datasets (Tables 1 and 2). The individual-level association with BMI was not apparent at the ecological level where ranking countries by median BMI revealed no clear gradient in median RHR, including when stratified by age group and sex (Figure S2). In SAB, however, test center-level RHR was positively associated with center-level BMI in both men and women (Supplementary Figure S8a) and this was also observed when using FMI as the measure of adiposity (Supplementary Figure S8b).

**Figure 2.**
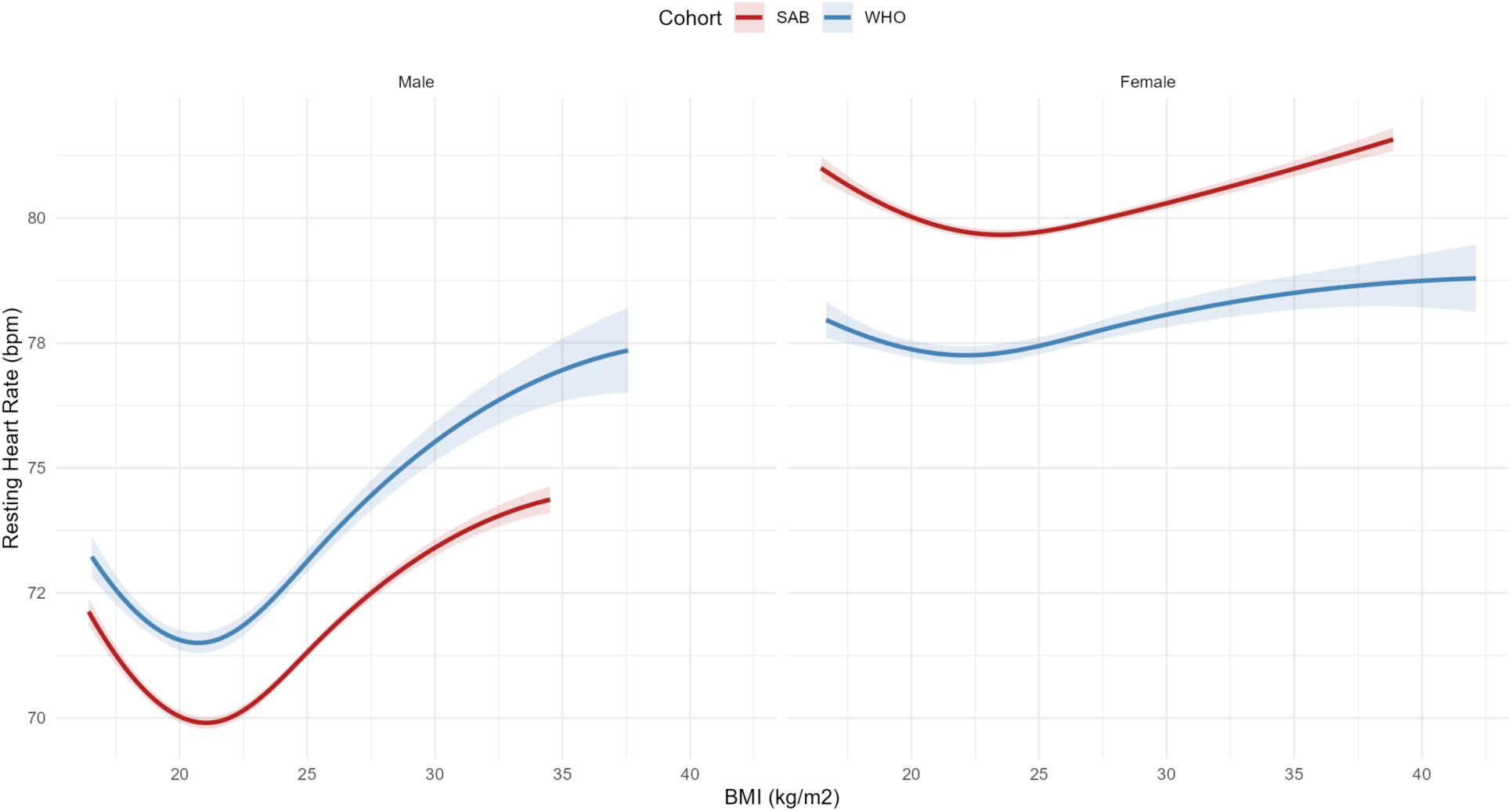
Sex-stratified cubic spline plots for the association between BMI and resting heart rate for WHO and SAB.

### Physical activity

Not meeting physical activity guidelines was associated with higher RHR in both sexes (WHO and SAB) (Tables 1 and 2). RHR was also inversely associated with device-measured physical activity in the SAB accelerometry subsample (Supplementary Figure S9).

### Lung function and disease status (SAB)

Better lung function, an alternative indicator of cardiorespiratory fitness, was associated with lower RHR (Supplementary Figure S9), whereas prevalent disease (hypertension, hypercholesterolemia, type 2 diabetes) was associated with higher RHR in both sexes (Table 2).

### Socio-economic status

Unemployment was associated with higher RHR in women but not men in WHO STEPS, whereas in SAB, it was associated with higher RHR in both men and women (Tables 1 and 2). Agricultural workers, cleaners, and unemployed participants generally had the lowest RHR.

Widowed status was associated with higher RHR in men but not in women in WHO STEPS. In SAB, however, widowed women, but not men, had lower RHR.

### Diet, smoking, and alcohol

RHR did not differ by fruit and vegetable intake (consuming at least 5 servings of fruits and vegetables a day). In SAB, smoking was associated with lower RHR (more so in women). Alcohol consumption was associated with higher RHR in men in both datasets, with the strongest association observed in frequent drinkers in SAB. In women, associations were slightly lower in both datasets (Tables 1 and 2).

### Variation across time

In 19 WHO countries with repeat survey data, RHR was stable or slightly declining, with a modest decrease only in men (−0.4 bpm per year) (Figure 3a). In SAB, RHR was higher mid-year (Table 2, Figure 3b), in both categorical (by month) and continuous (cosinor) analyses. Seasonal fluctuations were more pronounced for test regions further from the equator. Seasonal variation findings were supported by categorical analyses (Table 2).

**Figure 3.**
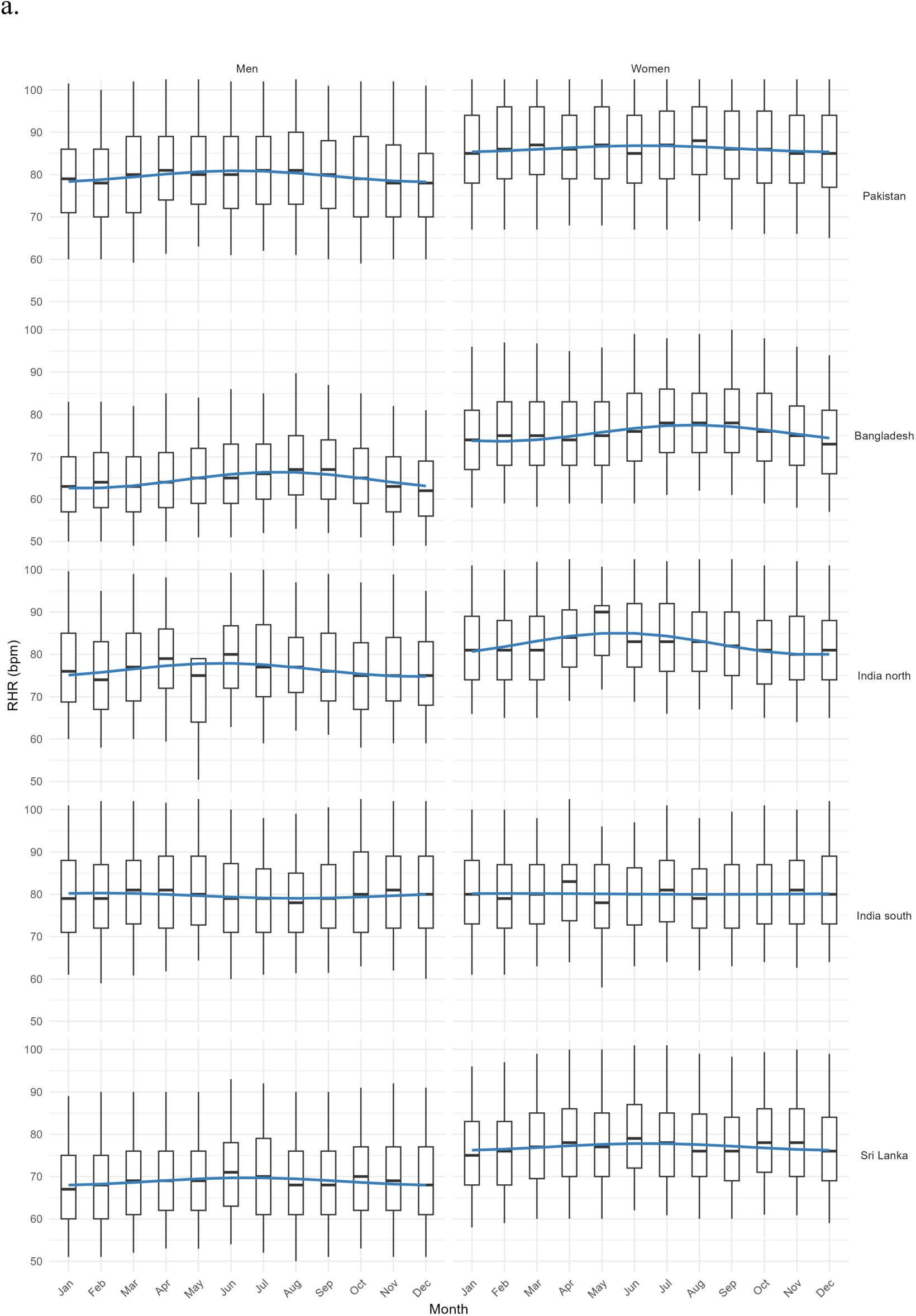
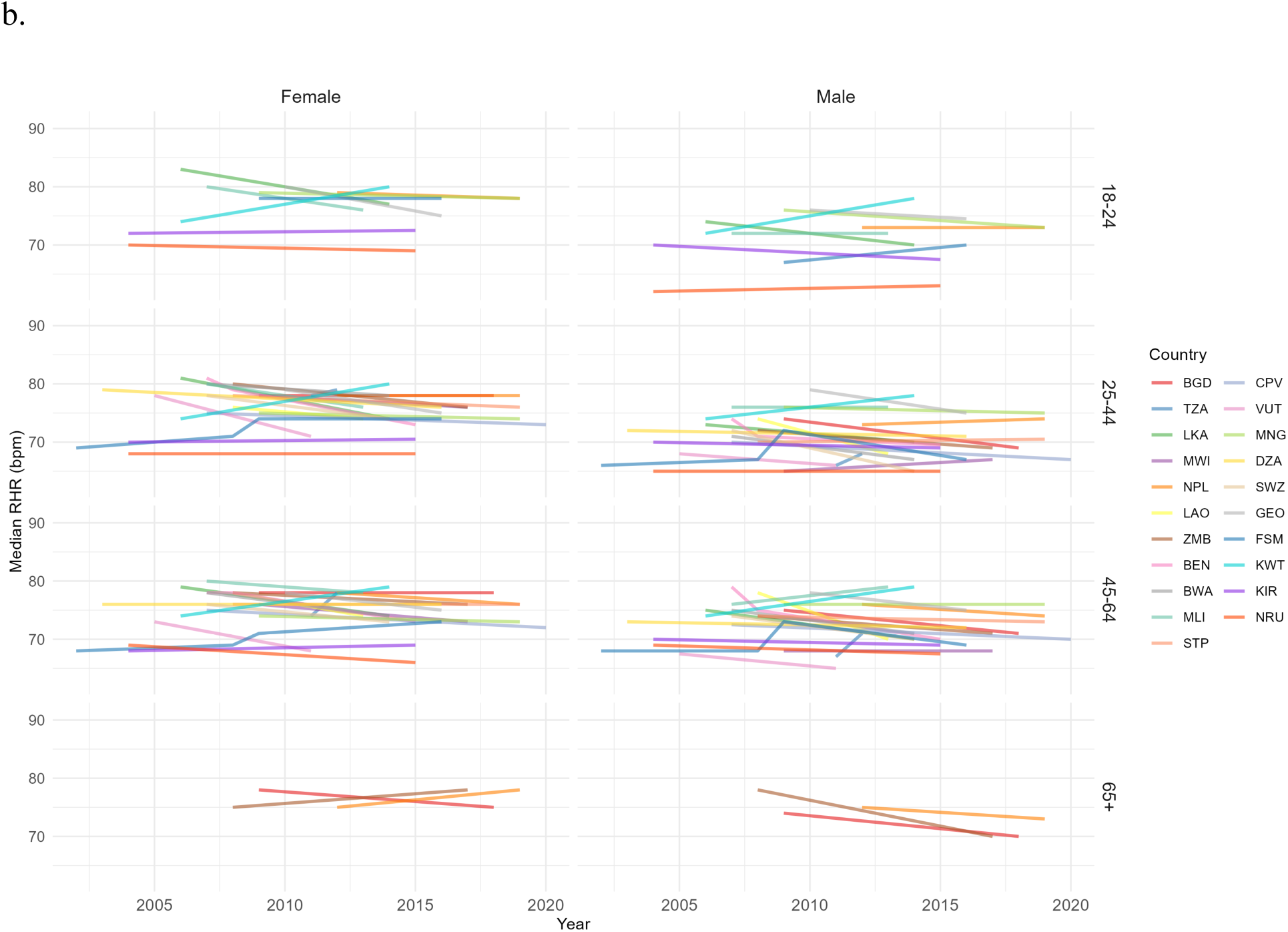
Sex-stratified line plots and box plots for the difference in resting heart rate over time. Panel a shows region-level (SAB) seasonal variation by month of data collection. Panel b shows country-level (WHO) time trends by survey year, stratified by age group.

### Contribution of resting heart rate to routine physical activity surveillance

To explore if RHR could add to the estimation of physical activity, we modeled how much variance in MVPA RHR could explain in SAB. Overall, RHR was a modest (negative) predictor of MVPA (Supplementary Table S1, S2). In the model with both RHR and GPAQ-based MVPA, both terms were predictors of device-based MVPA.

## DISCUSSION

### Main Findings

This study integrates WHO STEPS surveys and the South Asia Biobank to describe global and regional distributions of RHR and evaluate its potential utility in non-communicable disease surveillance. Age-related patterns diverged by sex, with RHR increasing and then declining with age in a curvilinear pattern in men across most regions, while in women RHR was steadily declining with age. Higher BMI and insufficient physical activity were universally associated with elevated RHR. Repeat cross-sectional survey results indicated long-term temporal stability with modest declines in men but seasonal variation was observed in non-equatorial regions. RHR added to the prediction of device-based physical activity, over and above that of self-reported physical activity. As RHR is already widely available in large population surveys and an easy-to-measure vital sign, our results suggest RHR may have utility in routine global health surveillance.

### Across the lifespan

In most regions, men exhibited a curvilinear age-RHR pattern, with higher RHR in midlife and lower values at older ages, whereas women showed a consistent decline. The rise in RHR in men to a midlife peak followed by decline mirrors patterns in two large wearable cohorts, 92,457 Fitbit users in the US and 4.8 million Fitbit users geolocated across 217 countries and territories (22,23). RHR patterns in women are less consistent. The US cohort showed a midlife peak while the global cohort showed a plateau or steady decline (22,23). Our findings extend these observations to population-based samples from low- and middle-income settings that are underrepresented in the literature.

The male curvilinear pattern likely reflects the competing physiological processes across the lifespan. Age-related reductions in parasympathetic tone elevate RHR through midlife, while selective survival may produce an apparent decline in the oldest age groups, given the established association between elevated RHR and cardiovascular mortality (18,24,25). The more linear decline observed in women may reflect sex-specific autonomic regulation, potentially mediated by hormonal influences on cardiovascular control (26). These divergent patterns suggest that sex differences in RHR reflect differences in cardiovascular ageing which may have implications for potential reference standards.

Within SAB, the association between RHR and age varied substantially across test regions in men. Only Sri Lankan men showed a monotonic decline but the relationship was inverse U-shaped in other countries. Women across all test regions showed monotonic declines, consistent with the WHO regional findings. As cardiometabolic disease patterns also vary across South Asia, in India for example diabetes prevalence ranges from 4.8% to 26.4% between states (27), healthy volunteer bias may also contribute to observed associations with varying response patterns in different test regions within SAB (20).

### Associations with modifiable factors

Higher BMI was associated with elevated RHR across both datasets, in both sexes, across all regions. In addition, insufficient physical activity was associated with higher RHR in both datasets, including when assessed by accelerometry in the SAB subsample, an assessment of physical behaviour that relates more strongly to health outcomes than self-reported behaviour (14). Prevalent metabolic disease (hypertension, hypercholesterolemia, and type 2 diabetes) was associated with higher RHR in SAB participants, consistent with prior evidence that RHR is associated with cardiometabolic risk (15,16). Taken together, these associations suggest that RHR captures meaningful biological information across diverse populations and measurement contexts.

### Temporality

In SAB, RHR was higher in June-August and this variation was more pronounced in test regions further from the equator. Similar results were observed in the largest study of RHR using wearables where summer–winter differences were 0.55 bpm in the Northern and 0.74 bpm in the Southern hemisphere (23). However, that study showed a peak in local winter, whereas SAB RHR peaked in summer. This may reflect heat stress or influence of time of day of readings (day vs night) (28). With that said, studies observing RHR differences across the year are inconsistent. Vukosavljević et al. found RHR to rise in unusually hot and cold temperatures while another study in Burkina Faso found no association between heat exposure and nocturnal heart rate (23,29). Irrespective of mechanism, an unbiased annual estimate of RHR requires repeat sampling across the year or statistical correction for season. WHO STEPS fieldwork is not not usually conducted to achieve full seasonal coverage, and date of measurement was also missing in most WHO data, hindering quantification of seasonal variation in this set (30).

In the WHO data with repeat surveys over longer time frames, RHR was broadly stable over time, with only a modest decline in men. This decline in men contrasts with recent literature documenting declining cardiorespiratory fitness globally that suggest the opposite (31). It is possible that differences in measurement protocols and participant selection across survey waves may partially explain this phenomenon (32,33). For example, participants in later survey years may be more or less rested than participants in earlier survey years due to different circumstances in the home where the measurement took place, and such biases over time could mask true changes in population RHR levels. Confirming genuine secular trends in RHR requires standardized measurement conditions that do not yet exist uniformly across national surveys, and it could be debated if survey designs have captured the true resting state of participants and consistently so over time (14). Although RHR is relatively robust to routine stressors shifting by less than 2% after hard training, it rises by around 6% during illness (34). As personal wearables with continuous heart rate monitoring capabilities are becoming more common, such data could support our findings with more robust measurements of the true resting heart rate level, for example during sleep, which is also the estimate of RHR that correlates more closely with cardiorespiratory fitness levels (14).

### Implications for Global Health Surveillance

The utility of RHR as a global surveillance tool relies on its accessibility and its biological validity. RHR is routinely captured alongside blood pressure in surveys like WHO STEPS, requires no additional equipment or specialist training, and is feasible in low-resource settings where VO_2_max testing is not (14). RHR is altered with physical training and is prospectively associated with health outcomes (18,35). Elevated RHR in high-risk regions, particularly in Pakistan where both RHR and adiposity were highest, underscores its utility. These patterns identify subpopulations at elevated metabolic disease risk where NCD burden is high and fitness data are scarce (12,36).

We also demonstrate the complementary value of RHR to self-reported physical activity which further strengthens the case for its use in routine surveillance. This was most evident in men. Device-measured activity in women was found to be poorly explained by all examined predictors in our study. This is possibly due to wrist-worn accelerometers detecting upper-body and domestic tasks that impose less sustained whole-body movement and cardiovascular load (37). The growing affordability of commercial wearables that continuously measure both physical activity and heart rate opens additional opportunities for personalized CVD risk stratification and community-based screening (38,39). Currently, however, most users are concentrated in high-income populations, with limited coverage in South Asia and Africa where NCD burden is high (23,40). Whilst wearable ownership is steadily increasing, surveys such as WHO STEPS and SAB are feasible avenues for capturing objective health indicators in populations underrepresented in the field.

### Strengths and Limitations

The strengths of this study include its large sample and global coverage. The use of population weighting enhances generalizability. Repeated cross-sectional surveys provides valuable insights into population-level time trends in RHR for routine health surveillance. Limitations include the cross-sectional observational design that limits causal inference, and differences between measurement protocols that may introduce heterogeneity and impact comparability. Studies using follow-up data and linkage to health outcomes in non-Western populations are needed to assert associations and strengthen causal interpretations.

### What is already known on this topic

- Cardiorespiratory fitness is strongly associated with CVD and mortality but remains understudied globally, particularly in non-Western populations.
- Resting heart rate is a feasible proxy indicator for cardiorespiratory fitness, routinely captured in national surveys, but its global epidemiology and surveillance potential have not been systematically described.

### What this study adds

- Globally, RHR generally ranges between 65 and 80 bpm
- Men and women show divergent age-related RHR trajectories, with a curvilinear pattern in men
- Higher BMI, insufficient physical activity, and prevalent metabolic disease are consistently associated with elevated RHR
- RHR can improve the prediction of objectively measured physical activity, over and above that of self-report

### How this study might affect research, practice or policy

- RHR offers a low-cost, accessible tool for fitness surveillance in low-resource settings.
- Sex-specific and region-specific RHR reference values may improve population-level risk stratification and inform targeted NCD interventions, particularly in South Asia.

## Acknowledgments

We would like to thank all survey participants for giving their time to the WHO STEPS and SAB. We are grateful to the many field teams of the WHO STEP Survey and the South Asia Biobank for their efforts in generating the datasets used here. We acknowledge the contribution of Dr. Fiona Bull and Dr. Leanne Riley from WHO head office, and the coordinators of the WHO surveys included in our study: Dr Djamila Nadir-Azirou (Algeria), Ms Tsagik Vardanyan and Ms Diana Andreasyan (Armenia), Dr Nabil Seyidov (Azerbaijan), Dr Delon Brennen (Bahamas), Prof Dr Baizid Khoorshid Riaz (Bangladesh), Dr Kenneth George (Barbados), Dr Vital Pisaryk (Belarus), Prof Dismand Stéphane Houinato (Benin), Dr Heluf G Medhin (Botswana), Dr Lubna binti Haji Abdul Razak (Brunei Darussalam), Dr Irenita Soares (Cabo Verde), Dr Sophal Oum (Cambodia), Prof Alfred K Njamnshi (Cameroon), Dr Anna Matthews (Cayman Islands), Dr Max Roger Koula (Central African Republic), Mr Mathias Roger Djidina (Chad), Dr Mohamed Msaidie (Comoros), Dr Kouamelan Doua (Côte d’Ivoire), Prof Longo Mbenza (Democratic Republic of the Congo), Romina Costa Beltrán (Ecuador), Dr. Goitom Mebrahtu Medhani (Eritrea), Dr. Lindiwe Tsabedze (Eswatini), Dr Isimeli Tukana (Fiji), Dr Comlan Pearl Nsie Obame (Gabon), Mr Omar Badjie (Gambia), Ms Lela Sturua (Georgia), Ms Agnes Banfield (Grenada), Prof Naby Moussa Baldé (Guinea), Dr Kavita Singh (Guyana), NCD Directorate at the Ministry of Health (Jordan), Dr Joseph Mwangi Kibachio (Kenya), Mrs Eretii Timeon (Kiribati), Dr Rihab Alwotayan (Kuwait), Dr Roza Djakypova (Kyrgyzstan), Dr Manithong Vonglokham (Lao People’s Democratic Republic), Ministry of Health (Lebanon), Dr Kabelo Mputsoe (Lesotho), Dr Peter L Clement (Liberia), Dr Omar Msalam (Libya), Dr Henri Fidele Marie Raharivohitra (Madagascar), Dr Kelias Msyamboza (Malawi), Mr Ubeydulla Thoufeeq (Maldives), Dr Diarra Nazoum (Mali), Prof Mohamed Lemine Ba (Mauritania), Mr Marcus Samo (Micronesia, Fed. Sts.), Dr Galina Obreja (Moldova, Republic of), Dr Narantuya Davaakhuu (Mongolia), Dr Hicham El Berri (Morocco), Dr Carla MT Da Silva Matos (Mozambique), Dr Tint Swe Latt (Myanmar), Mr Don Kadir (Nauru), Dr Meghnath Dhimal and Mr Bihungum Bista (Nepal), Dr Moussa Habibou (Niger), Ms Grizelda Mokoia (Niue), Mr Alypio Nyandwi (Rwanda), Dr Alina Jaime (Saint Lucia), Dr Lazaro Sousa (São Tomé and Principe), Dr Mohamed Samai (Sierra Leone), Dr Geoffrey Kenilorea and Mrs Nevalyn Laesango (Solomon Islands), Dr Lakshmi C Somatunga (Sri Lanka), Dr Nazik Hassan Babiker (Sudan), Mr Saifudinov Safar (Tajikistan), Dr Joao Martins (Timor-Leste), Dr Happy Kokou Agoudavi (Togo), Dr Silivia Tavite (Tokelau), Mr Sioape Kupu (Tonga), Felise Manoa Afasene (Tuvalu), Dr Gerald Mutungi (Uganda), Dr Mary Mayige (United Republic of Tanzania), Dr Miskiya Mohamed (United Republic of Tanzania (Zanzibar)), Dr Maria Jose Rodriguez (Uruguay), Douglas Ngwele (Vanuatu), Dr Tran Quoc Bao (Viet Nam), Mr Tareq Ramahi (West Bank and Gaza Strip), Dr Namasiku Siyumbwa (Zambia). We also thank the WHO and national governments and donors that funded the individual surveys included in this study. AA and SB designed the analyses and drafted the manuscript. All authors reviewed / edited the content and take full responsibility for the submitted version. Analysis scripts were developed with assistance from ChatGPT for initial syntax generation and debugging. All code was manually reviewed, tested for correctness, validated against raw data and revised as necessary. The final scripts will be made publicly available on GitHub upon acceptance.

SAB was funded by the National Institute for Health Research (NIHR) (16/136/68 and 132960) using UK aid from the UK Government to support global health research. ADA was supported by the Kuwait Foundation for the Advancement of the Sciences. TG and SB were supported by the UK Medical Research Council (MC_UU_00006/4) and the NIHR Biomedical Research Centre in Cambridge (NIHR203312). LF was supported by the NIHR (NIHR133205) and the Medical Research Council (grant number UKRI1478).

The funders had no role in study design, data collection, analysis, interpretation, or reporting.

## Contributors

ADA, TG and SB designed the analysis. ADA analysed the data. ADA, TG and SB drafted the manuscript. MaL, SA, PK, VJ, AK, KIK, MM, RMA, JCC, WX, MH and SB contributed to the design of the South Asia Biobank. MaL, SA, PK, VJ, AK, KIK, MM, RMA, JCC, WX, PA, GP, SM, AB, LDS, MC, AT, MeL, MH, AA and TS collected data. The South Asia Biobank Study Group contributed to the design, implementation and data collection of the South Asia Biobank. All named authors contibuted to the interpetation of findings and reviewed the manuscript and approved the final version. The WHO STEPS survey country coordinators listed in the Acknowledgements led data collection in their countries and are collectively credited as non-author contributors. ADA and SB are guarantors: they accept full responsibility for the work and the conduct of the study, had access to the data, and controlled the decision to publish.

## AI Declaration

Analysis scripts were developed with assistance from Claude for initial syntax generation and debugging. All code was manually reviewed, tested for correctness, validated against raw data and revised.

## Patient and public involvement

Patients and/or the public were not involved in the design, conduct, reporting or dissemination plans of this research.

## Patient consent for publication

Not applicable.

## Ethics approval

The South Asia Biobank was approved by the Imperial College London Research Ethics Committee (reference 18IC4698) and by local institutional review boards in each participating country. Each WHO STEPS survey was approved by the appropriate national ethics body. Participants gave informed consent to participate in the study before taking part.

## Data availability statement

Data are available upon reasonable request. WHO STEPS survey data are available on request from the WHO NCD Microdata Repository (https://extranet.who.int/ncdsmicrodata/index.php/home). South Asia Biobank data are available to researchers on request to the study Steering Committee.

## APPENDIX

**Figure S1.**
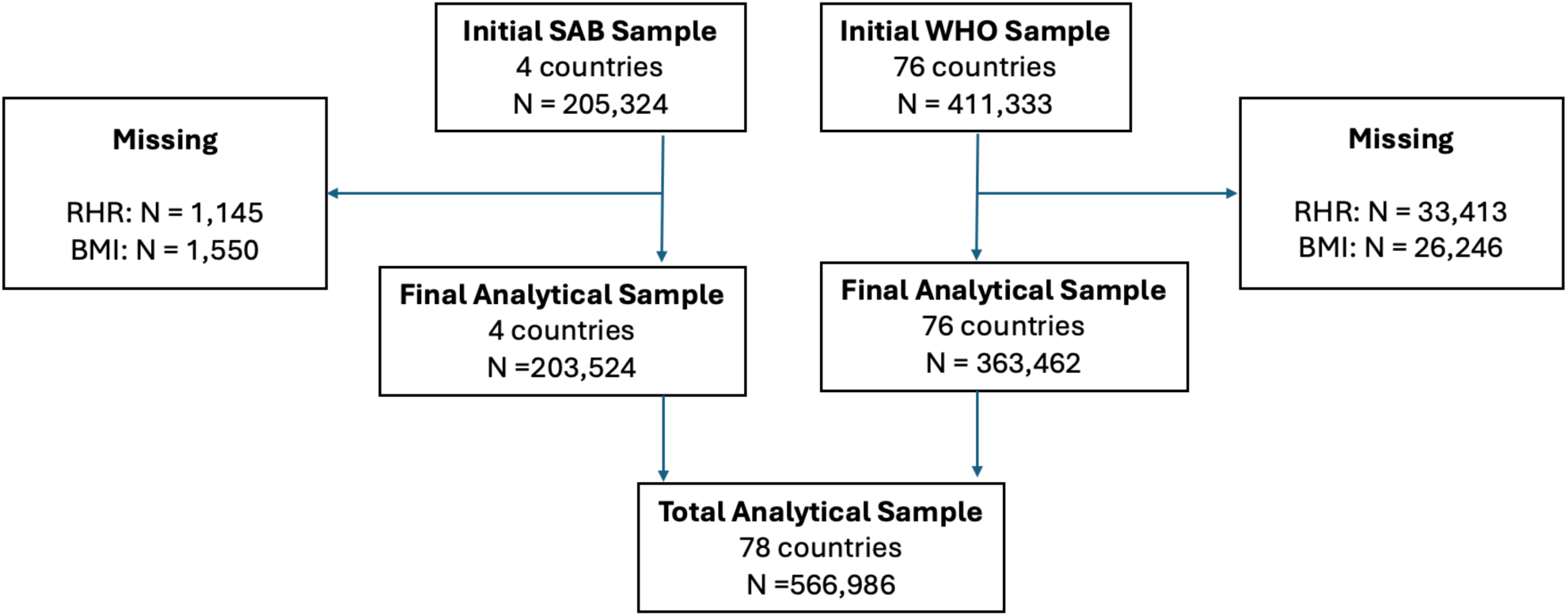
Flowchart for participants included in the SAB and WHO analyses, respectively.

**Figure S2.**
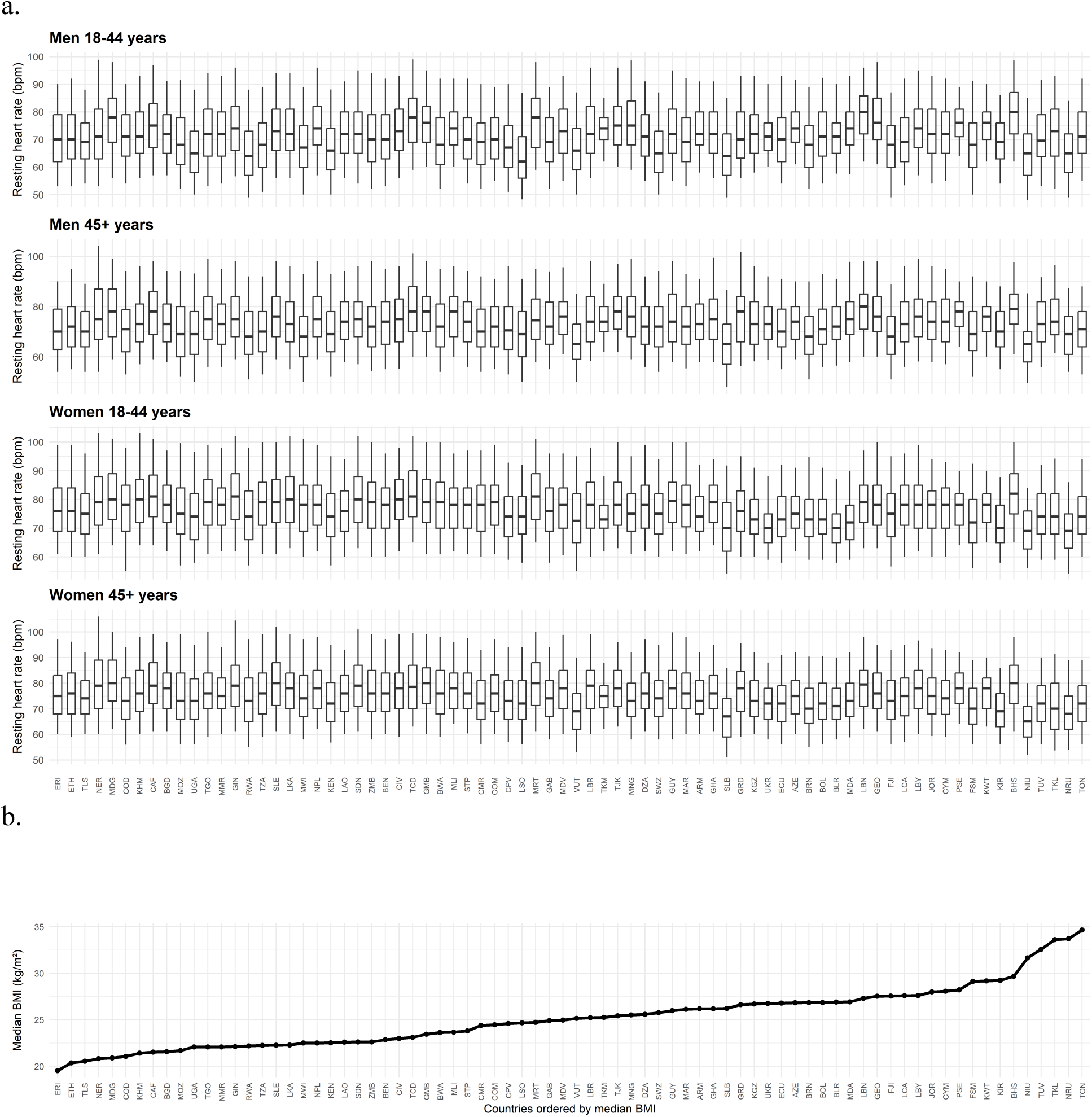
Note: Median BMI is pooled across both men and women.

**Figure S3.**
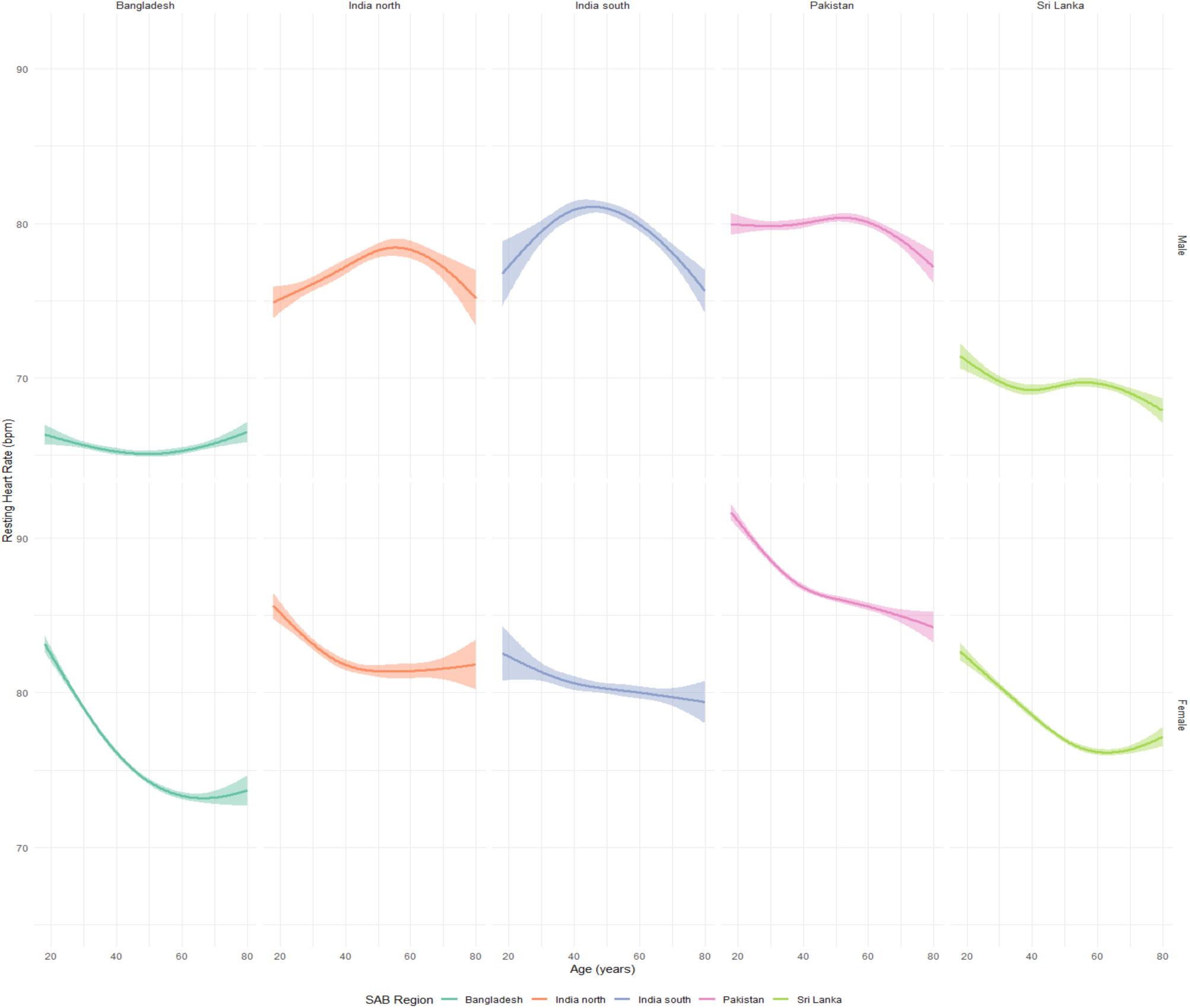
Sex-stratified cubic spline models for the association between age and resting heart rate by SAB region.

**Figure S4.**
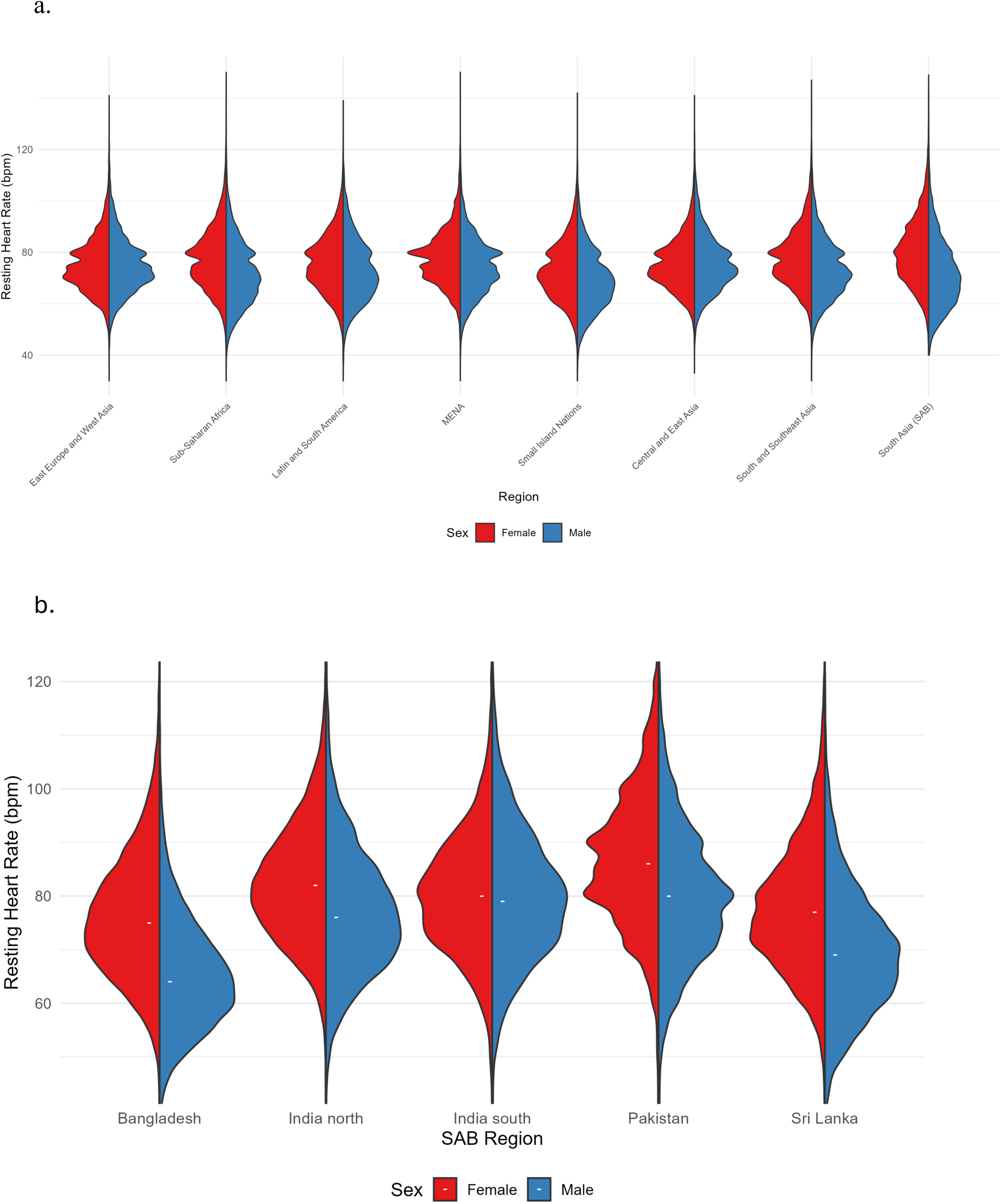
Sex-stratified violin plots for RHR distribution by world region (WHO) and test region (SAB).

**Figure S5.**
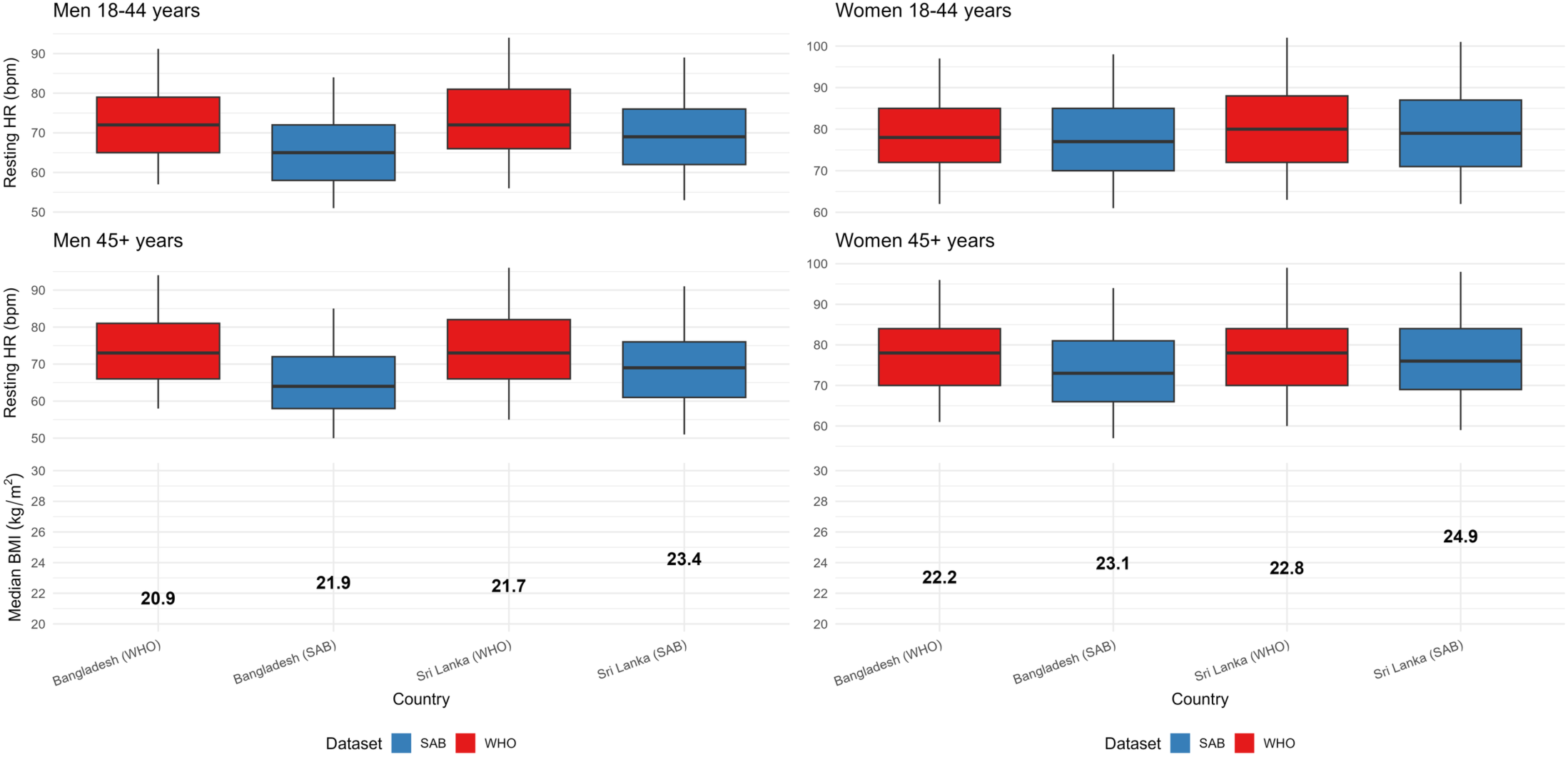
Comparison of SAB and WHO measurements in Bangladesh and Sri Lanka samples, stratified by sex and age group RHR medians exhibited sex-specific patterns when comparing SAB and WHO datasets from Bangladesh and Sri Lanka with significant differences confirmed by Wilcoxon tests in all age and sex strata. For men, SAB medians were generally lower than those in WHO, ranging from 64–66 bpm in Bangladesh (versus 68–73 bpm in WHO) and 68–71 bpm in Sri Lanka (versus 69– 70 bpm in WHO). For women, SAB medians were higher, ranging from 73–81 bpm in Bangladesh (versus 74–76 bpm in WHO) and 76–81 bpm in Sri Lanka (versus 73–77 bpm in WHO). These patterns may reflect true population differences, potentially influenced by temporal shifts (SAB 2019–2023 vs. WHO 2009–2018 for Bangladesh, 2014 for Sri Lanka) and differences in methodology.

**Figure S6.**
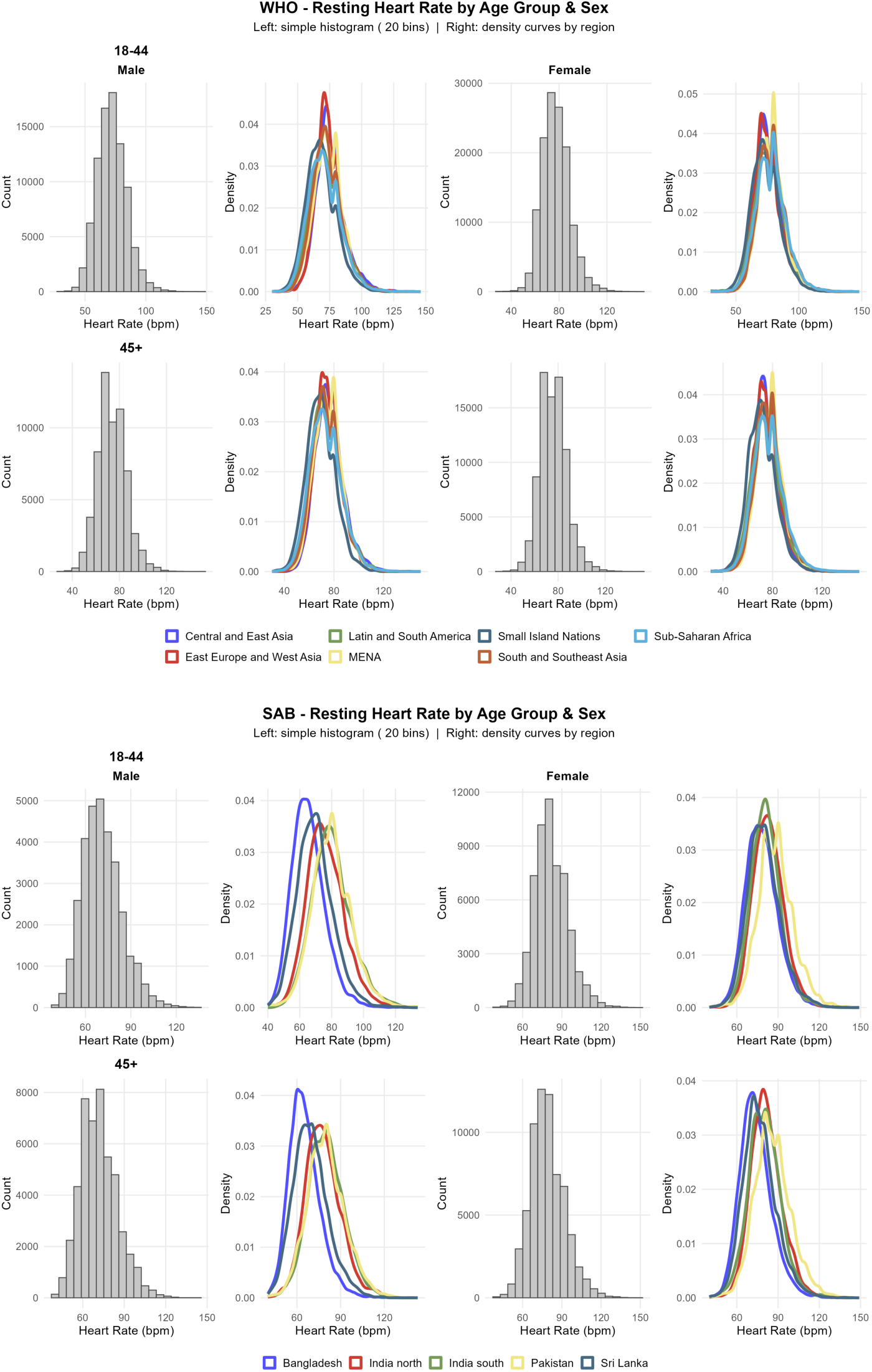
Histograms and density plots for the distribution of resting heart rate across WHO world regions and SAB test regions, stratified by sex and age group.

**Figure S7.**
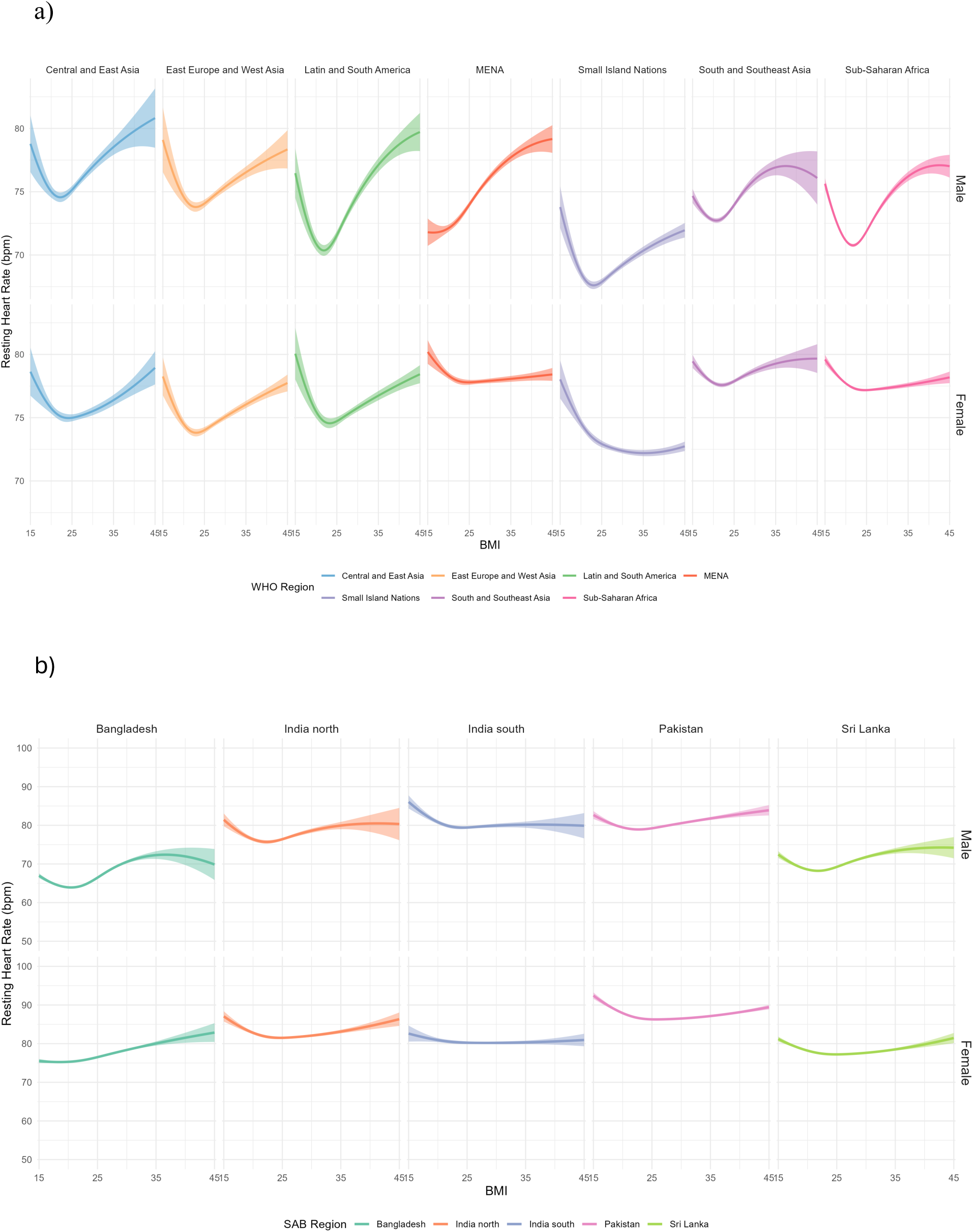
Sex-stratified cubic spline models for the association between BMI and resting heart rate by a) WHO region and b) SAB test region.

**Figure S8:**
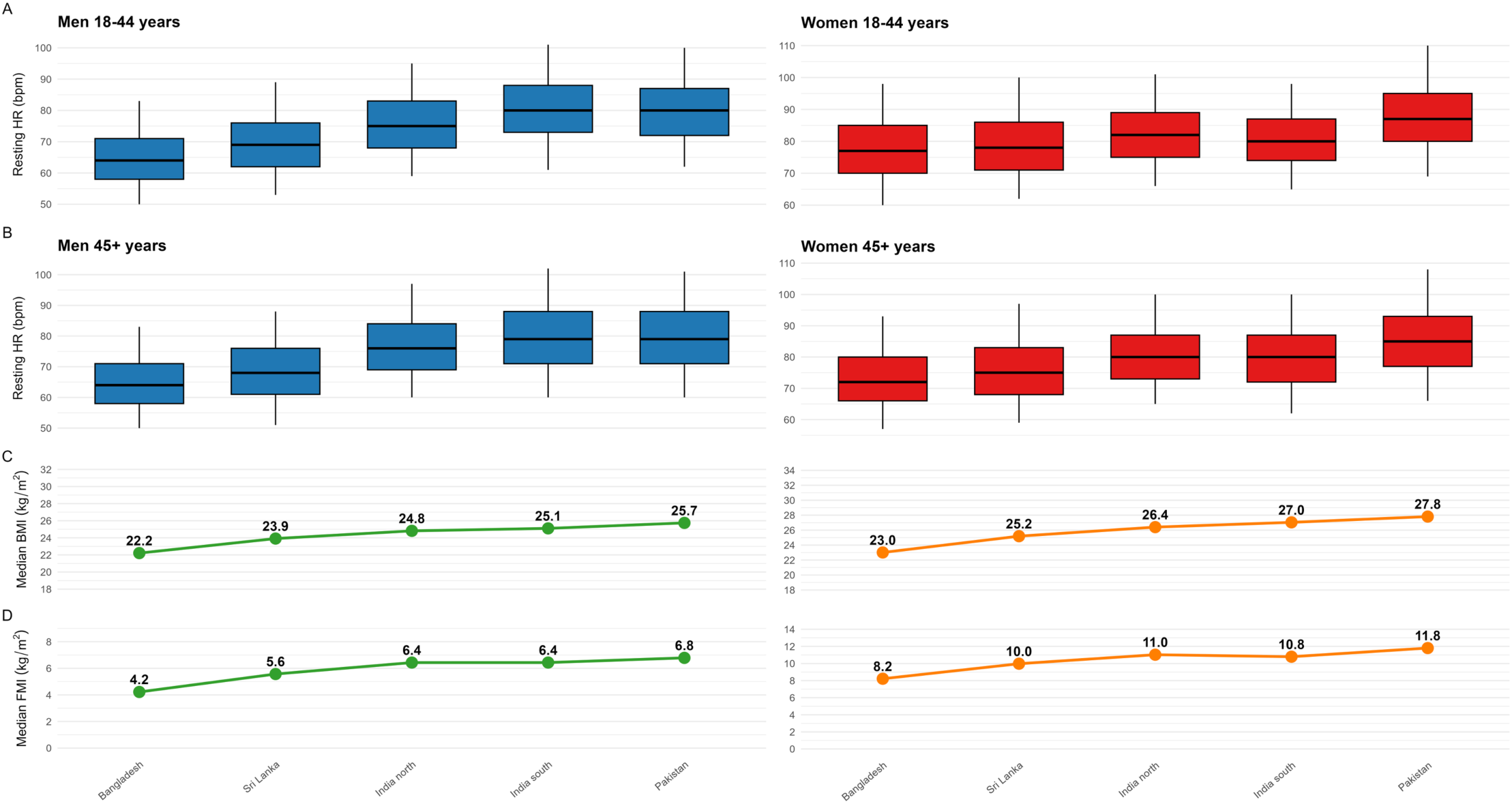
Sex and age-stratified boxplots of resting heart rate in each SAB test region ordered by median BMI and FMI.

**Figure S9.**
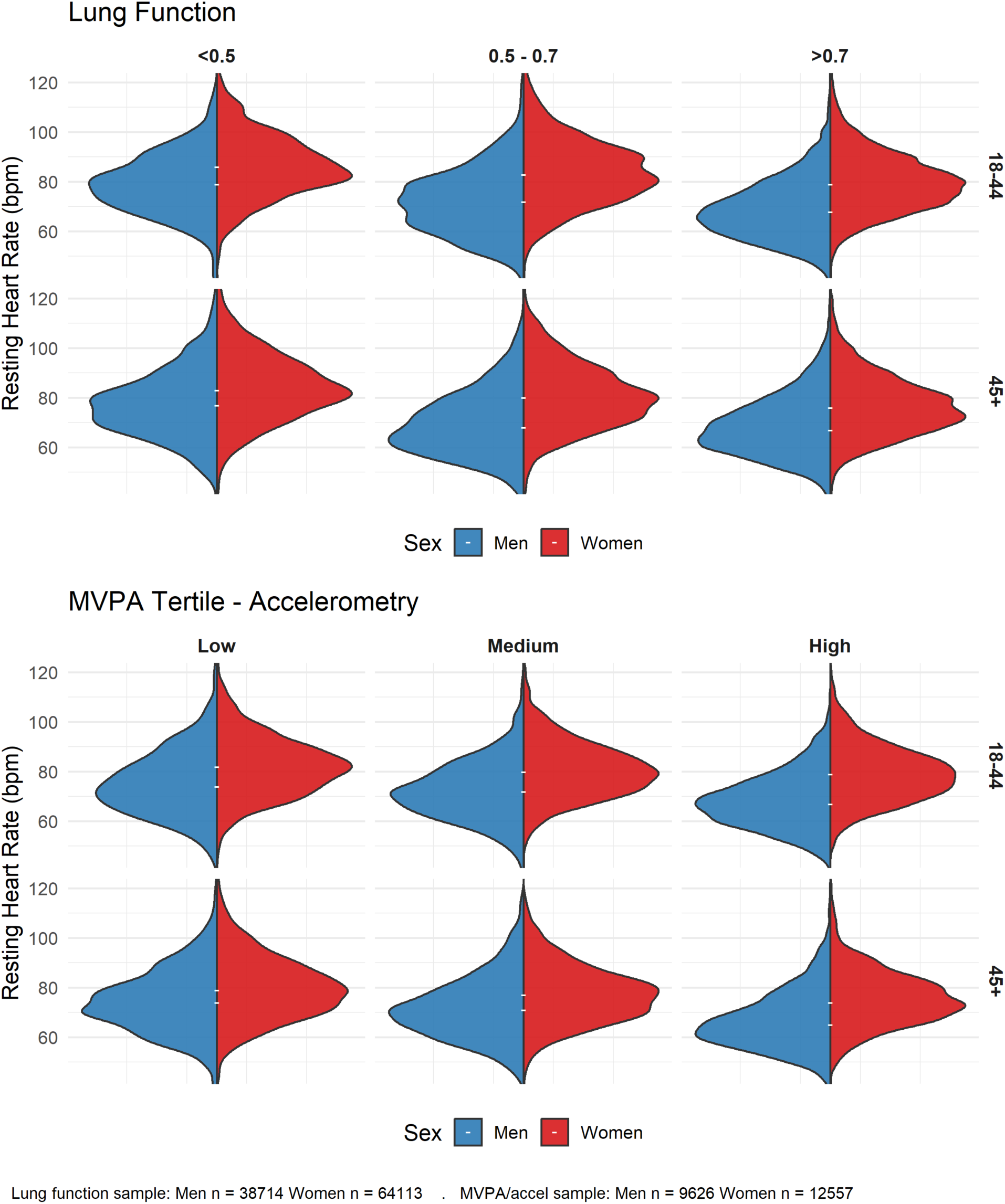
Violin plots for the distribution of resting heart rate across lung function categories and device-based MVPA tertiles stratified by sex and age group (SAB).

**Supplementary Material S1.** Comparison of included and excluded participants (Excel File)

**Supplementary Material S2:** Univariate and multivariable adjusted association analysis estimates of resting heart rate (WHO and SAB) (Excel file)

**Supplementary Table S1.**
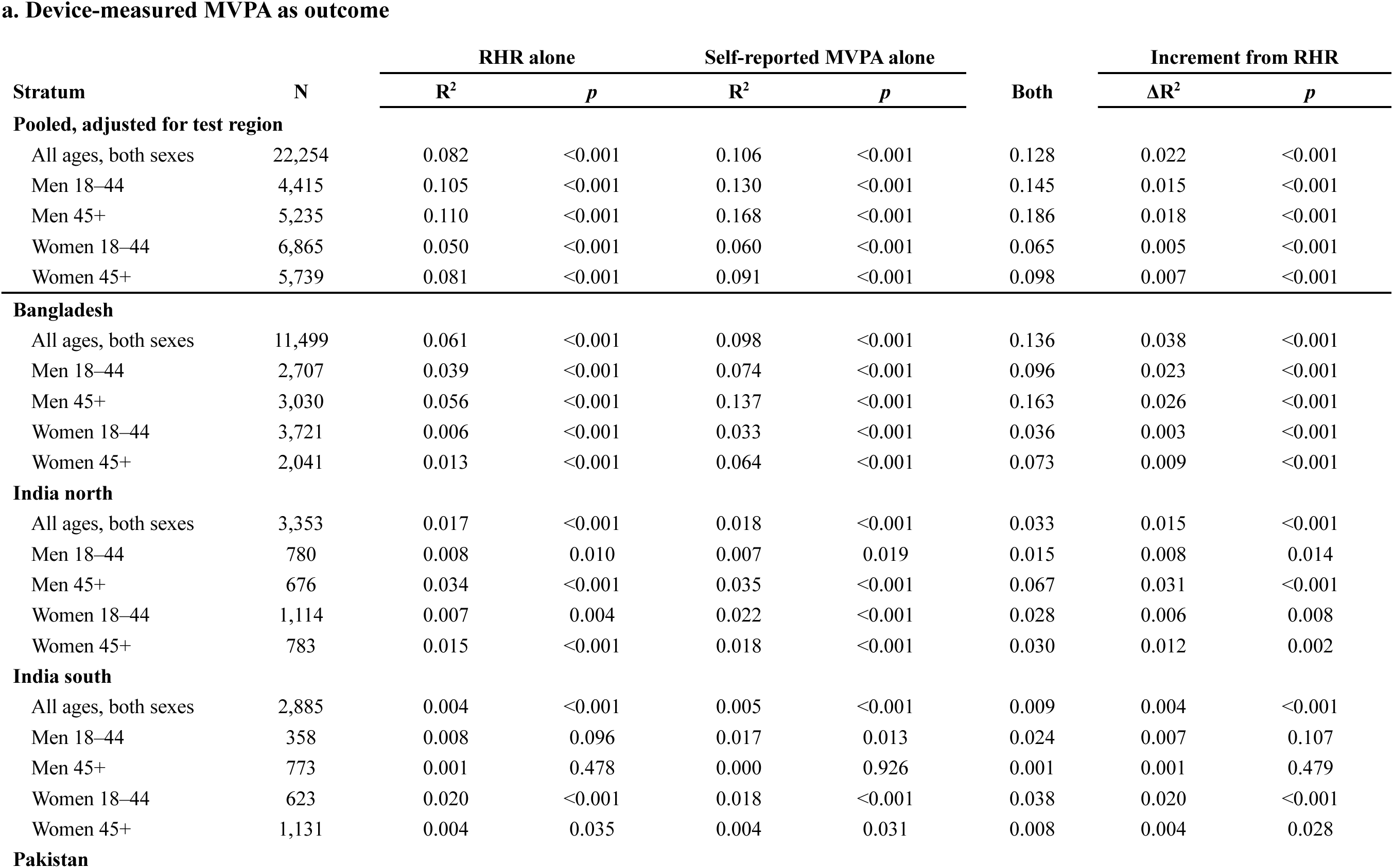
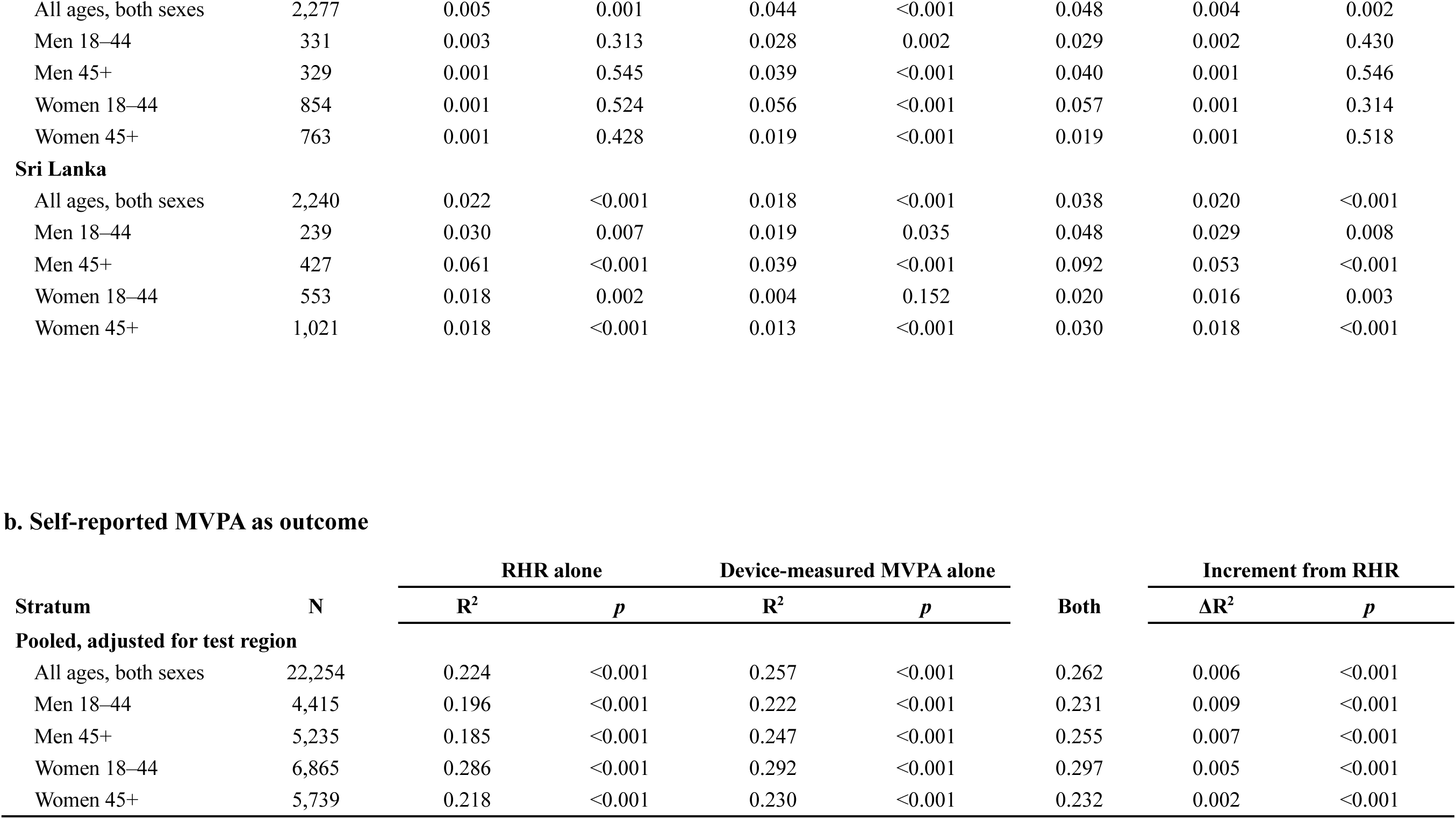
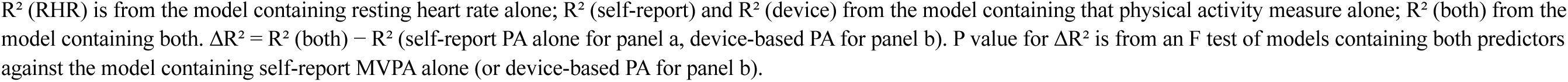
Proportion of variance in device-based and self-report physical activity by resting heart rate in SAB.

| **a. Device-measured MVPA as outcome** |  |  |  |  |  |  |  |  |
|---|---|---|---|---|---|---|---|---|
|  |  | **RHR alone** |  | **Self-reported MVPA alone** |  |  | **Increment from RHR** |  |
| **Stratum** | **N** | **R<sup>2</sup>** | ***p*** | **R<sup>2</sup>** | ***p*** | **Both** | **ΔR<sup>2</sup>** | ***p*** |
| **Pooled, adjusted for test region** |  |  |  |  |  |  |  |  |
| All ages, both sexes | 22,254 | 0.082 | <0.001 | 0.106 | <0.001 | 0.128 | 0.022 | <0.001 |
| Men 18–44 | 4,415 | 0.105 | <0.001 | 0.130 | <0.001 | 0.145 | 0.015 | <0.001 |
| Men 45+ | 5,235 | 0.110 | <0.001 | 0.168 | <0.001 | 0.186 | 0.018 | <0.001 |
| Women 18–44 | 6,865 | 0.050 | <0.001 | 0.060 | <0.001 | 0.065 | 0.005 | <0.001 |
| Women 45+ | 5,739 | 0.081 | <0.001 | 0.091 | <0.001 | 0.098 | 0.007 | <0.001 |
| **Bangladesh** |  |  |  |  |  |  |  |  |
| All ages, both sexes | 11,499 | 0.061 | <0.001 | 0.098 | <0.001 | 0.136 | 0.038 | <0.001 |
| Men 18–44 | 2,707 | 0.039 | <0.001 | 0.074 | <0.001 | 0.096 | 0.023 | <0.001 |
| Men 45+ | 3,030 | 0.056 | <0.001 | 0.137 | <0.001 | 0.163 | 0.026 | <0.001 |
| Women 18–44 | 3,721 | 0.006 | <0.001 | 0.033 | <0.001 | 0.036 | 0.003 | <0.001 |
| Women 45+ | 2,041 | 0.013 | <0.001 | 0.064 | <0.001 | 0.073 | 0.009 | <0.001 |
| **India north** |  |  |  |  |  |  |  |  |
| All ages, both sexes | 3,353 | 0.017 | <0.001 | 0.018 | <0.001 | 0.033 | 0.015 | <0.001 |
| Men 18–44 | 780 | 0.008 | 0.010 | 0.007 | 0.019 | 0.015 | 0.008 | 0.014 |
| Men 45+ | 676 | 0.034 | <0.001 | 0.035 | <0.001 | 0.067 | 0.031 | <0.001 |
| Women 18–44 | 1,114 | 0.007 | 0.004 | 0.022 | <0.001 | 0.028 | 0.006 | 0.008 |
| Women 45+ | 783 | 0.015 | <0.001 | 0.018 | <0.001 | 0.030 | 0.012 | 0.002 |
| **India south** |  |  |  |  |  |  |  |  |
| All ages, both sexes | 2,885 | 0.004 | <0.001 | 0.005 | <0.001 | 0.009 | 0.004 | <0.001 |
| Men 18–44 | 358 | 0.008 | 0.096 | 0.017 | 0.013 | 0.024 | 0.007 | 0.107 |
| Men 45+ | 773 | 0.001 | 0.478 | 0.000 | 0.926 | 0.001 | 0.001 | 0.479 |
| Women 18–44 | 623 | 0.020 | <0.001 | 0.018 | <0.001 | 0.038 | 0.020 | <0.001 |
| Women 45+ | 1,131 | 0.004 | 0.035 | 0.004 | 0.031 | 0.008 | 0.004 | 0.028 |
| **Pakistan** |  |  |  |  |  |  |  |  |
| All ages, both sexes | 2,277 | 0.005 | 0.001 | 0.044 | <0.001 | 0.048 | 0.004 | 0.002 |
| Men 18–44 | 331 | 0.003 | 0.313 | 0.028 | 0.002 | 0.029 | 0.002 | 0.430 |
| Men 45+ | 329 | 0.001 | 0.545 | 0.039 | <0.001 | 0.040 | 0.001 | 0.546 |
| Women 18–44 | 854 | 0.001 | 0.524 | 0.056 | <0.001 | 0.057 | 0.001 | 0.314 |
| Women 45+ | 763 | 0.001 | 0.428 | 0.019 | <0.001 | 0.019 | 0.001 | 0.518 |
| **Sri Lanka** |  |  |  |  |  |  |  |  |
| All ages, both sexes | 2,240 | 0.022 | <0.001 | 0.018 | <0.001 | 0.038 | 0.020 | <0.001 |
| Men 18–44 | 239 | 0.030 | 0.007 | 0.019 | 0.035 | 0.048 | 0.029 | 0.008 |
| Men 45+ | 427 | 0.061 | <0.001 | 0.039 | <0.001 | 0.092 | 0.053 | <0.001 |
| Women 18–44 | 553 | 0.018 | 0.002 | 0.004 | 0.152 | 0.020 | 0.016 | 0.003 |
| Women 45+ | 1,021 | 0.018 | <0.001 | 0.013 | <0.001 | 0.030 | 0.018 | <0.001 |
| **b. Self-reported MVPA as outcome** |  |  |  |  |  |  |  |  |
|  |  | **RHR alone** |  | **Device-measured MVPA alone** |  |  | **Increment from RHR** |  |
| **Stratum** | **N** | **R<sup>2</sup>** | ***p*** | **R<sup>2</sup>** | ***p*** | **Both** | **ΔR<sup>2</sup>** | ***p*** |
| **Pooled, adjusted for test region** |  |  |  |  |  |  |  |  |
| All ages, both sexes | 22,254 | 0.224 | <0.001 | 0.257 | <0.001 | 0.262 | 0.006 | <0.001 |
| Men 18–44 | 4,415 | 0.196 | <0.001 | 0.222 | <0.001 | 0.231 | 0.009 | <0.001 |
| Men 45+ | 5,235 | 0.185 | <0.001 | 0.247 | <0.001 | 0.255 | 0.007 | <0.001 |
| Women 18–44 | 6,865 | 0.286 | <0.001 | 0.292 | <0.001 | 0.297 | 0.005 | <0.001 |
| Women 45+ | 5,739 | 0.218 | <0.001 | 0.230 | <0.001 | 0.232 | 0.002 | <0.001 |

R<sup>2</sup> (RHR) is from the model containing resting heart rate alone; R<sup>2</sup> (self-report) and R<sup>2</sup> (device) from the model containing that physical activity measure alone; R<sup>2</sup> (both) from the model containing both. ΔR<sup>2</sup> = R<sup>2</sup> (both) − R<sup>2</sup> (self-report PA alone for panel a, device-based PA for panel b). P value for ΔR<sup>2</sup> is from an F test of models containing both predictors against the model containing self-report MVPA alone (or device-based PA for panel b).

**Supplementary Table S2.**
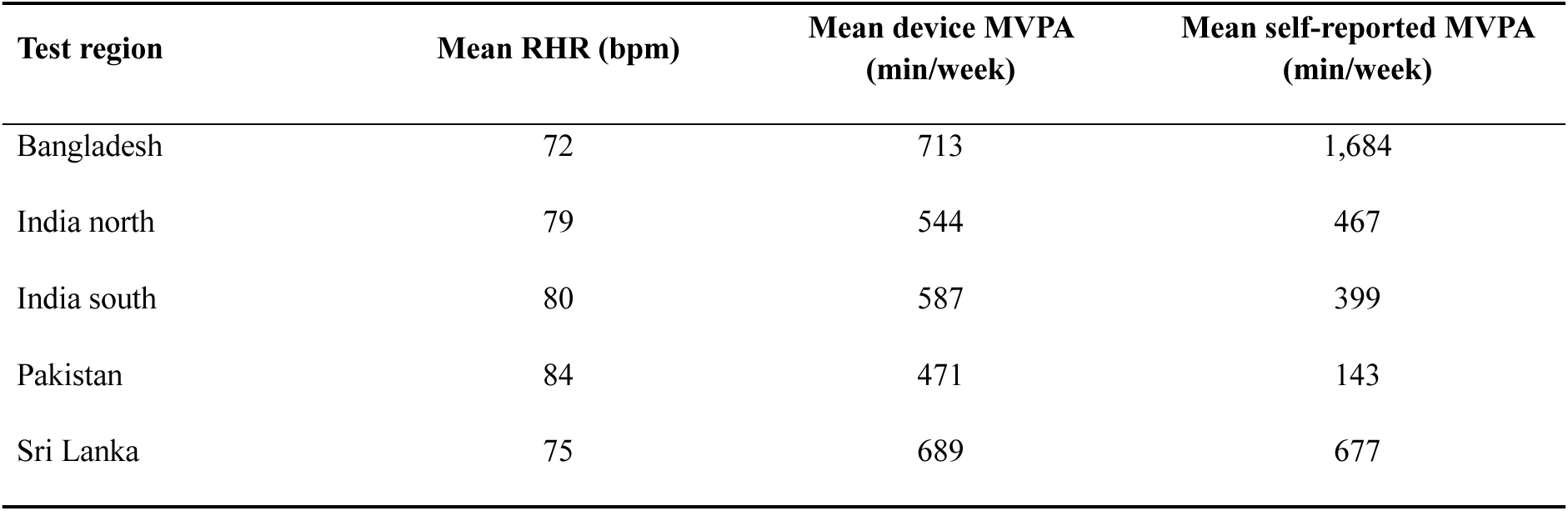
Test region means.

| **Test region** | **Mean RHR (bpm)** | **Mean device MVPA (min/week)** | **Mean self-reported MVPA (min/week)** |
|---|---|---|---|
| Bangladesh | 72 | 713 | 1,684 |
| India north | 79 | 544 | 467 |
| India south | 80 | 587 | 399 |
| Pakistan | 84 | 471 | 143 |
| Sri Lanka | 75 | 689 | 677 |

## REFERENCES

1. Lang JJ, Prince SA, Merucci K, Cadenas-Sanchez C, Chaput JP, Fraser BJ, et al. Cardiorespiratory fitness is a strong and consistent predictor of morbidity and mortality among adults: an overview of meta-analyses representing over 20.9 million observations from 199 unique cohort studies. Br J Sports Med. 2024 May 1;58(10):556–66. doi:10.1136/bjsports-2023-107849

2. Ng M, Gakidou E, Lo J, Abate YH, Abbafati C, Abbas N, et al. Global, regional, and national prevalence of adult overweight and obesity, 1990–2021, with forecasts to 2050: a forecasting study for the Global Burden of Disease Study 2021. Lancet. 2025 Mar;405(10481):813–38. doi:10.1016/S0140-6736(25)00355-1

3. Zeiher J, Ombrellaro KJ, Perumal N, Keil T, Mensink GBM, Finger JD. Correlates and determinants of cardiorespiratory fitness in adults: a systematic review. Sports Med Open. 2019 Dec;5(1):39. doi:10.1186/s40798-019-0211-2

4. Gonzales TI, Westgate K, Hollidge S, Lindsay T, Wijndaele K, Forouhi NG, et al. Descriptive epidemiology of cardiorespiratory fitness in UK adults: the Fenland Study. Med Sci Sports Exerc. 2023 Mar;55(3):507–16. doi:10.1249/MSS.0000000000003068

5. Weeldreyer NR, De Guzman JC, Paterson C, Allen JD, Gaesser GA, Angadi SS. Cardiorespiratory fitness, body mass index and mortality: a systematic review and meta-analysis. Br J Sports Med. 2024 Nov 13;bjsports-2024-108748. doi:10.1136/bjsports-2024-108748

6. Arango CM, Parra DC, Gómez LF, Lema L, Lobelo F, Ekelund U. Screen time, cardiorespiratory fitness and adiposity among school-age children from Monteria, Colombia. J Sci Med Sport. 2014 Sep;17(5):491–5. doi:10.1016/j.jsams.2013.09.010

7. Christensen DL, Jørgensen SW, Koch LS, Nordsborg NB, Sironga J, Ramaiya KL, et al. Directly measured aerobic fitness in male Maasai of Tanzania. Am J Hum Biol. 2022 Apr;34(4):e23674. doi:10.1002/ajhb.23674

8. Prioreschi A, Brage S, Westgate K, Norris SA, Micklesfield LK. Cardiorespiratory fitness levels and associations with physical activity and body composition in young South African adults from Soweto. BMC Public Health. 2017 Dec;17(1):301. doi:10.1186/s12889-017-4212-0

9. Christensen DL, Faurholt-Jepsen D, Boit MK, Mwaniki DL, Kilonzo B, Tetens I, et al. Cardiorespiratory fitness and physical activity in Luo, Kamba, and Maasai of rural Kenya. Am J Hum Biol. 2012 Nov;24(6):723–9. doi:10.1002/ajhb.22303

10. Hingorjo MR, Zehra S, Hasan Z, Qureshi MA. Cardiorespiratory fitness and its association with adiposity indices in young adults. Pak J Med Sci. 2017 Jun 2;33(3). doi:10.12669/pjms.333.12294

11. Christensen DL, Alcalá-Sánchez I, Leal-Berumen I, Conchas-Ramirez M, Brage S. Physical activity, cardio-respiratory fitness, and metabolic traits in rural Mexican Tarahumara. Am J Hum Biol. 2012 Jul;24(4):558–61. doi:10.1002/ajhb.22239

12. Ghouri N, Purves D, McConnachie A, Wilson J, Gill JMR, Sattar N. Lower cardiorespiratory fitness contributes to increased insulin resistance and fasting glycaemia in middle-aged South Asian compared with European men living in the UK. Diabetologia. 2013 Oct;56(10):2238–49. doi:10.1007/s00125-013-2969-y

13. Gonzales TI, Westgate K, Strain T, Hollidge S, Jeon J, Christensen DL, et al. Cardiorespiratory fitness assessment using risk-stratified exercise testing and dose– response relationships with disease outcomes. Sci Rep. 2021 Jul 28;11(1):15315. doi:10.1038/s41598-021-94768-3

14. Gonzales TI, Jeon JY, Lindsay T, Westgate K, Perez-Pozuelo I, Hollidge S, et al. Resting heart rate is a population-level biomarker of cardiorespiratory fitness: the Fenland Study. PLoS One. 2023 May 11;18(5):e0285272. doi:10.1371/journal.pone.0285272

15. Kelley E, Imboden MT, Harber MP, Finch H, Kaminsky LA, Whaley MH. Cardiorespiratory fitness is inversely associated with clustering of metabolic syndrome risk factors: the Ball State Adult Fitness Program Longitudinal Lifestyle Study. Mayo Clin Proc Innov Qual Outcomes. 2018 Jun;2(2):155–64. doi:10.1016/j.mayocpiqo.2018.03.001

16. Sloan RA. Estimated cardiorespiratory fitness and metabolic risks. Int J Environ Res Public Health. 2024 May 16;21(5):635. doi:10.3390/ijerph21050635

17. Gutierrez-Martinez L, Brellenthin AG, Lefferts EC, Lee DC, Sui X, Lavie CJ, et al. Resting heart rate and risk of cancer mortality. Cancer Epidemiol Biomarkers Prev. 2021 Jun 1;30(6):1072–8. doi:10.1158/1055-9965.EPI-20-1731

18. Raisi-Estabragh Z, Cooper J, Judge R, Khanji MY, Munroe PB, Cooper C, et al. Age, sex and disease-specific associations between resting heart rate and cardiovascular mortality in the UK BIOBANK. PLoS One. 2020 May 29;15(5):e0233898. doi:10.1371/journal.pone.0233898

19. Riley L, Guthold R, Cowan M, Savin S, Bhatti L, Armstrong T, et al. The World Health Organization STEPwise approach to noncommunicable disease risk-factor surveillance: methods, challenges, and opportunities. Am J Public Health. 2016 Jan;106(1):74–8. doi:10.2105/AJPH.2015.302962

20. Song P, Gupta A, Goon IY, Hasan M, Mahmood S, Pradeepa R, et al. Data resource profile: understanding the patterns and determinants of health in South Asians—the South Asia Biobank. Int J Epidemiol. 2021 Jul 9;50(3):717–718e. doi:10.1093/ije/dyab029

21. Backman H, Vanfleteren LEGW, Mannino DM, Ekström M. Severity of airflow obstruction based on FEV1/FVC versus FEV1 percent predicted in the general U.S. population. Am J Respir Crit Care Med. 2024 Dec 1;210(11):1308–16. doi:10.1164/rccm.202310-1773OC

22. Quer G, Gouda P, Galarnyk M, Topol EJ, Steinhubl SR. Inter-and intraindividual variability in daily resting heart rate and its associations with age, sex, sleep, BMI, and time of year: retrospective, longitudinal cohort study of 92,457 adults. PLoS One. 2020 Feb 5;15(2):e0227709. doi:10.1371/journal.pone.0227709

23. Vukosavljević K, Schönfeldt A, Phillips J, Cho Y, Zhu T, Patel S, et al. Global geographic, seasonal, environmental and demographic drivers of variations in longitudinal cardiac physiology in 5 million Fitbit users. Research Square [Preprint]. Version 1. 2026 Sep 14 [cited 2026 Oct 23]. doi:10.21203/rs.3.rs-10785318/v1

24. Shaffer F, Ginsberg JP. An overview of heart rate variability metrics and norms. Front Public Health. 2017 Sep 28;5:258. doi:10.3389/fpubh.2017.00258

25. Jensen MT. Resting heart rate and relation to disease and longevity: past, present and future. Scand J Clin Lab Invest. 2019 Feb 17;79(1–2):108–16. doi:10.1080/00365513.2019.1566567

26. Koenig J, Thayer JF. Sex differences in healthy human heart rate variability: a meta-analysis. Neurosci Biobehav Rev. 2016 May;64:288–310. doi:10.1016/j.neubiorev.2016.03.007

27. Anjana RM, Unnikrishnan R, Deepa M, Pradeepa R, Tandon N, Das AK, et al. Metabolic non-communicable disease health report of India: the ICMR-INDIAB national cross-sectional study (ICMR-INDIAB-17). Lancet Diabetes Endocrinol. 2023 Jul;11(7):474–89. doi:10.1016/S2213-8587(23)00119-5

28. Crandall CG, Wilson TE. Human cardiovascular responses to passive heat stress. Compr Physiol. 2015 Jan;5(1):17–43. doi:10.1002/cphy.c140015

29. Koch M, Matzke I, Huhn S, Sié A, Boudo V, Compaoré G, et al. Assessing the effect of extreme weather on population health using consumer-grade wearables in rural Burkina Faso: observational panel study. JMIR Mhealth Uhealth. 2023 Nov 8;11:e46980. doi:10.2196/46980

30. Brage S, Assah F, Msyamboza KP. Quantifying population levels of physical activity in Africa using wearable sensors: implications for global physical activity surveillance. BMJ Open Sport Exerc Med. 2020 Oct;6(1):e000941. doi:10.1136/bmjsem-2020-000941

31. Hills AP, Jayasinghe S, Arena R, Byrne NM. Global status of cardiorespiratory fitness and physical activity – are we improving or getting worse? Prog Cardiovasc Dis. 2024 Mar;83:16–22. doi:10.1016/j.pcad.2024.02.008

32. Kaminsky LA, Myers J, Arena R. Determining cardiorespiratory fitness with precision: compendium of findings from the FRIEND Registry. Prog Cardiovasc Dis. 2019 Jan;62(1):76–82. doi:10.1016/j.pcad.2018.10.003

33. Lequeux B, Uzan C, Rehman MB. Does resting heart rate measured by the physician reflect the patient’s true resting heart rate? White-coat heart rate. Indian Heart J. 2018 Jan;70(1):93–8. doi:10.1016/j.ihj.2017.07.015

34. Altini M, Plews D. What is behind changes in resting heart rate and heart rate variability? A large-scale analysis of longitudinal measurements acquired in free-living. Sensors. 2021 Nov 27;21(23):7932. doi:10.3390/s21237932

35. Reimers AK, Knapp G, Reimers CD. Effects of exercise on the resting heart rate: a systematic review and meta-analysis of interventional studies. J Clin Med. 2018 Dec 1;7(12):503. doi:10.3390/jcm7120503

36. Kaminsky LA, Arena R, Ellingsen Ø, Harber MP, Myers J, Ozemek C, et al. Cardiorespiratory fitness and cardiovascular disease – the past, present, and future. Prog Cardiovasc Dis. 2019 Mar;62(2):86–93. doi:10.1016/j.pcad.2019.01.002

37. Liu F, Wanigatunga AA, Schrack JA. Assessment of physical activity in adults using wrist accelerometers. Epidemiol Rev. 2021 Dec 30;43(1):65–93. doi:10.1093/epirev/mxab004

38. Feng KY, Short SA, Saeb S, Carroll MK, Olivier CB, Simard EP, et al. Resting heart rate and associations with clinical measures from the Project Baseline Health Study: observational study. J Med Internet Res. 2024 Dec 20;26:e60493. doi:10.2196/60493

39. Dunn J, Kidzinski L, Runge R, Witt D, Hicks JL, Schüssler-Fiorenza Rose SM, et al. Wearable sensors enable personalized predictions of clinical laboratory measurements. Nat Med. 2021 Jun;27(6):1105–12. doi:10.1038/s41591-021-01339-0

40. Strain T, Wijndaele K, Pearce M, Brage S. Considerations for the use of consumer-grade wearables and smartphones in population surveillance of physical activity. J Meas Phys Behav. 2022;5(1):8–14. doi:10.1123/jmpb.2021-0046

